# Childhood conduct problems, potential snares in adolescence and problematic substance use in Brazil

**DOI:** 10.1101/2024.09.18.24313884

**Authors:** Fauve Stocker, Jon Heron, Matthew Hickman, Fernando C. Wehrmeister, Helen Gonçalves, Ana Maria B. Menezes, Joseph Murray, Gemma Hammerton

**Affiliations:** Population Health Sciences, Bristol Medical School, University of Bristol, Bristol, UK; Medical Research Council Integrative Epidemiology Unit at the University of Bristol, Population Health Sciences, Bristol Medical School, University of Bristol, Bristol, UK and the Centre for Academic Mental Health, Population Health Sciences, Bristol Medical School, University of Bristol, Bristol, UK; Health Protection Research Unit in Behavioural Science and Evaluation (HPRU BSE) at the University of Bristol, Population Health Sciences, Bristol Medical School, Bristol, UK; Postgraduate Program in Epidemiology, Universidade Federal de Pelotas, Pelotas, Brazil; Human Development and Violence Research Centre (DOVE), Federal University of Pelotas, Pelotas, Brazil

**Author notes:** contributed equally to this work (Joseph Murray, Gemma Hammerton). **Correspondence to** Gemma Hammerton, PhD, Population Health Sciences, University of Bristol, Canynge Hall, Bristol, UK, BS8 2PS.

**Keywords:** conduct problems, substance use, counterfactual mediation, Brazil, 1993 Pelotas Birth Cohort

## Abstract

**Objective:** Examine whether the relationship between childhood conduct problems and substance use in adulthood is explained through potential snares (police arrest, gang membership, school non-completion) in a Brazilian population-based birth cohort.

**Method:** Data were analyzed from 4,599 young people from the 1993 Pelotas Birth Cohort in Brazil. The exposure was conduct problems at age 11 years. Outcomes included hazardous alcohol consumption and illicit drug use (age 22 years). Mediators included police arrest (by age 18 years), gang membership (ages 18 and 22 years), and school non-completion (by age 22 years). We performed counterfactual mediation using the parametric g-computation formula to estimate the indirect effect via all three mediators simultaneously.

**Results:** After adjusting for confounders (including comorbidity), conduct problems were weakly associated with police arrest [OR (95% CI)=1.45 (0.97, 2.16)] and school non-completion [OR (95% CI)=1.46 (1.22, 1.74)], but not with gang membership. Police arrest and gang membership were associated with illicit drug use [OR (95% CI)=3.84 (2.46, 5.99); OR (95% CI)=7.78 (4.30, 14.10), respectively] and with hazardous alcohol use [OR (95% CI)=1.60 (1.08, 2.38); OR (95% CI)=1.88 (1.07, 3.30)]. However, there was no association between school non-completion and either outcome after confounder adjustment. There was little evidence for an indirect effect of conduct problems on hazardous alcohol use and illicit drug use via all three mediators after confounder adjustment.

**Conclusion:** The indirect effects of childhood conduct problems on substance use in early adulthood via police arrest, gang membership, and school non-completion were very small, after accounting for confounders and comorbidity.

## Introduction

Conduct problems refer to behaviors related to conduct disorder (norm-breaking behaviors and violations of the rights of others) and behaviors related to oppositional defiant disorder (noncompliant, angry and defiant behaviours). In Brazil, conduct problems are almost twice as prevalent as in high-income countries (HIC), with an average prevalence of 21% compared to 13% in the UK and 11% in the US (measured using the Strengths and Difficulties Questionnaire)^1^, with a higher prevalence even after accounting for measurement differences^2^. There is robust evidence that childhood conduct problems are associated with various adverse outcomes in early adulthood, including problem behaviors such as alcohol and drug misuse^3^. Although the majority of evidence comes from HIC, a previous study comparing a British and Brazilian birth cohort found that associations for childhood conduct problems with hazardous alcohol use and illicit drug use in early adulthood were stronger in Brazil compared to the UK^2^.

Problematic substance use places a significant burden on society as it is linked with alcohol dependency, road traffic accidents, injury, violence, and crime^4^. To reduce problematic substance use in Brazil, it is important to understand factors that are on the causal pathway from childhood conduct problems. An influential developmental taxonomy published by Moffitt in 1993 introduced the concept of snares that can trap adolescents into long-term problematic behaviors beyond an age when most otherwise desist from these behaviors^5^. Previous literature has suggested that interrupted education, gang membership and contact with the criminal justice system can act as snares trapping some adolescents into persistent problem behavior, including problematic substance use, when desistance is normative^3,6–8^.

Very little research has examined the importance of potential snares in low- and middle-income countries (LMIC). This is particularly important to address in countries, such as Brazil, with a higher prevalence of both conduct problems^1^ and potential snares such as gang membership and school non-completion^9,10^, compared to HIC. In LMIC settings, the processes for the association between conduct problems and later problematic substance use might differ substantially from HIC due to fewer supportive systems and different social contexts for children with conduct problems^1,11^.

Considering school non-completion as a possible “snare” linking conduct problems and later substance use, in Brazil, repetitive grade retention is a very common practice^12^ and is a strong predictor of dropping out of school^13^. Longitudinal data show Brazilian children repeating a school grade by age 11 had increased odds of elevated conduct problems into mid-adolescence^14^, and those with an externalising disorder in adolescence had higher odds of grade repetition three years later^15^. The association between childhood conduct problems and school non-completion is also well-documented in HIC^16–20^. However, it is not clear if there is a causal relationship between conduct problems and school non-completion, with some studies suggesting it is confounded by inattention^19^. There is evidence that school non-completion is associated with problematic behaviors in early adulthood, such as substance use^21,22^, and completing school by age 22 years was associated with lower odds of violent and non-violent crime, even after accounting for school grade repetitions in Brazil^23^.

Considering another possible “snare” following child conduct problems, youth gangs are more prevalent in Brazil than in most HIC and may offer a form of social capital and protection to adolescents who have failed to comply with societal norms^9,24^. Excessive alcohol, drug use and interpersonal violence are common among gang members and becoming a member increases the chances of early initiation and frequent consumption of alcohol and illicit drugs^25^. There is evidence that childhood conduct problems are a risk factor for gang membership in both HIC and LMIC^9,24,26^. Finally, contact with the criminal justice system can be a consequence of childhood conduct problems, given the strong links between childhood conduct problems and later crime in LMIC^2,27^. According to labelling theory, stigmatising effects of obtaining a criminal record are internalised by young people, resulting in self-stigma, low self-efficacy and self-esteem, increasing susceptibility to long-term problematic alcohol and drug use^28^.

One of the challenges for understanding relationships between child conduct problems, potential snares (such as gang membership) and later substance use, is the possibility of bidirectional relationships and shared or distinct sets of confounders for each relationship. Addressing these challenges requires data from a study where young people are followed longitudinally across childhood, adolescence, and early adulthood, with potential confounders assessed across multiple domains. In the current study, we examine whether potential snares in late adolescence (school non-completion, police arrest, and gang membership) mediate the association between childhood conduct problems and hazardous alcohol use and illicit drug use in early adulthood using a population-based, prospective birth cohort in Brazil: the 1993 Pelotas Birth Cohort.

In a previous study using the 1993 Pelotas Birth Cohort, conduct problems at age 11 years were associated with hazardous alcohol use and illicit drug use at age 22 years, after accounting for sociodemographic, individual, peer, family, and community confounders^2^. In this study, we hypothesise that associations between conduct problems at age 11 years and hazardous alcohol use and illicit drug use at age 22 years will be partly explained through potential snares between age 18 and 22 years (including school non-completion, police arrest, and gang membership). We hypothesise that potential baseline and intermediate confounders will weaken these indirect effects, particularly when accounting for childhood hyperactivity on the associations for conduct problems^2,19,29,30^.

## Methods

### Sample

The 1993 Pelotas Birth Cohort Study is an ongoing population-based study designed to investigate the effects of a wide range of influences on health and development. Pelotas is a city located in the extreme south of Brazil, with an estimated population of 345,000 inhabitants, 93% of whom live in the urban area. All births occurring in the five maternity clinics in the town were monitored in 1993 (99% of births in Pelotas occurred in hospital). For the 5,265 children born alive, only 16 mothers could not be interviewed or refused to participate in the study. The 5,249 newborns, whose mothers lived in the urban area, were included in the cohort (81 were either twins or triplets). The detailed methodology of this study can be found elsewhere^31,32^. During the perinatal study, mothers were interviewed to collect demographic, health and socioeconomic information about the family. Follow-up visits were conducted in 2004–2005 (age 11; *N* = 4,452 mothers; retention rate of 87.5%, after accounting for *N* = 141 deaths), 2008 (age 15; 4,349 mothers; retention rate of 85.7%, after accounting for *N* = 147 deaths), 2011-2012 (age 18; 4,106 mothers; retention rate of 81.4%, after accounting for *N* = 164 deaths), and 2015-2016 (age 22; *N* = 3,810 young people; retention rate of 76.3%, after accounting for *N* = 193 deaths)^31,33^. Study data at age 22 years was collected and managed using REDCap electronic data capture tools^34^. The perinatal study and each follow-up were approved by the Research Ethics Committee of the Federal University of Pelotas School of Medicine. After being informed of the details of the study, participants signed a term of informed consent.

### Measures

#### Exposure: Conduct problems at age 11 years

Conduct problems were assessed at age 11 during an interview with the primary caregiver (usually mothers) using the Strengths and Difficulties Questionnaire (SDQ)^35^. The SDQ is a screening questionnaire for children’s mental health problems that occurred in the previous six months, and has been validated in Brazil^36,37^ using independently diagnosed psychiatric disorders. Five items form the conduct problems subscale, including: ‘often has temper tantrums or hot tempers’, ‘often fights with other children or bullies them’, ‘often lies or cheats’, ‘steals from home, school or elsewhere’ and ‘generally obedient, usually does what adults request’. Responses for each item are ‘Not true’, ‘Somewhat true’ and ‘Certainly true’. The Item ‘generally obedient’ was reverse coded. All items were summed together (range 0 to 10) and the scale was dichotomised to create a binary exposure of low conduct problems (less than four) versus high conduct problems (four or more) according to the cut point on the newer 4-band categorisation (https://www.sdqinfo.org/py/sdqinfo/c0.py).

#### Outcomes: Hazardous alcohol use and illicit drug use at age 22 years

The 10-item self-report Alcohol Use Disorder Identification Test (AUDIT^38^) was used to assess hazardous alcohol consumption at age 22 years. The AUDIT is a brief screening tool aiming to detect risky drinking and risk for alcohol dependence with high validity and reliability^39^ and has been validated using diagnostic interview, physical examinations, and laboratory testing^40^. The AUDIT was dichotomised at a cut-point of eight and treated as a binary variable. The cut-point of eight or more represents hazardous levels of drinking and has been validated in Brazil^41^ using psychiatric diagnoses of alcohol use disorders.

Illicit drug use was assessed using the same self-report questionnaire at age 22 years which included questions about lifetime use of cannabis, cocaine, crack, amphetamine-type stimulants, nitrous oxide or other inhalants, hallucinogens, opioids, and other injected illegal drugs. A binary variable was created representing current drug use with “no” representing ‘never used’ (61%), ‘I just tried it’ (17%), and ‘I used to use it but I don’t anymore’ (8%) and “yes” representing ‘I use it occasionally’ (7%), ‘I only use it on weekends’ (1%), and ‘I use it every day or almost every day’ (5%).

#### Mediators: Police arrest (age 18), gang membership (age 18-22), and school non-completion (22 years)

Police arrest was assessed by self-report at the age of 18, asking if the young person had ever been arrested or detained (“yes” if ever arrested or detained and “no” otherwise).

Gang membership was assessed by self-report at ages 18 and 22 years with questions asking whether the participant had been a gang member in the previous year. Both time points were combined into a binary variable representing gang membership at either time point due to the low prevalence of gang membership (approximately 1% at both time points).

School non-completion (assessed at age 22) was a dichotomous variable indicating if the young person has finished high school (passed all school grades) by age 22 (“yes” if not completed school and “no” if completed). In Brazil, young people typically finish school by age 17, assuming they do not fail any grades; however, a student may continue studying into adulthood until they complete all grades^23^.

#### Baseline and intermediate confounders

Baseline confounders and intermediate confounders were drawn from five domains (sociodemographic, individual, peer, family, and community) that have previously been identified as key factors influencing a young person’s behavior (see Supplement 1, available online). Baseline confounders included sex, a score of sociodemographic (e.g., low maternal education) and health (e.g., mother smoking in pregnancy) risk factors for conduct problems (assessed by maternal report or observation during the perinatal period), maternal depression, the quality of the father-child relationship, the quality of the mother-child relationship, parental alcohol consumption, parental smoking status, parental separation, and neighbourhood safety (all assessed at child age 11 years). Intermediate confounders included peer deviance (assessed at child age 11 years), frequency of cigarette smoking and alcohol consumption, and peer drug use (all assessed at child age 15 years).

In secondary analyses, hyperactivity problems (at age 11 years) were considered as an additional baseline confounder and emotional problems (at age 15 years) were considered as an additional intermediate confounder.

### Statistical analyses

Prior to conducting any statistical analysis, we constructed a Directed Acyclic Graph (DAG) to present hypothesised causal pathways based on previously published articles (see Figure 1). All statistical analyses were carried out using Stata version 17^42^. First, we performed descriptive statistics to report characteristics of the population for the whole sample. Second, we conducted unadjusted and adjusted logistic regressions in stages to examine exposure-mediator and mediator-outcome associations. Third, we performed a counterfactual approach to mediation using the parametric g-computation formula to estimate indirect effects using binary mediators and common binary outcomes, and incorporate intermediate confounders^43^. The package -gformula-in Stata^44^ was used to estimate the total causal effect, natural direct effect and natural indirect effect via all three mediators simultaneously. All mediators were included simultaneously as we did not consider them to be independent. However, given that we were not able to confidently specify a causal direction between, we were only able to estimate one indirect effect via all three mediators together. -gformula-uses Monte Carlo simulations to simulate the mediator, outcome, and intermediate confounders under each hypothetical “counter to the fact” scenario. Standard errors were estimated using 50 bootstrap samples and normal-based 95% confidence intervals were calculated. More detail on the statistical analyses can be found in Supplement 2, available online.

**Figure 1.**
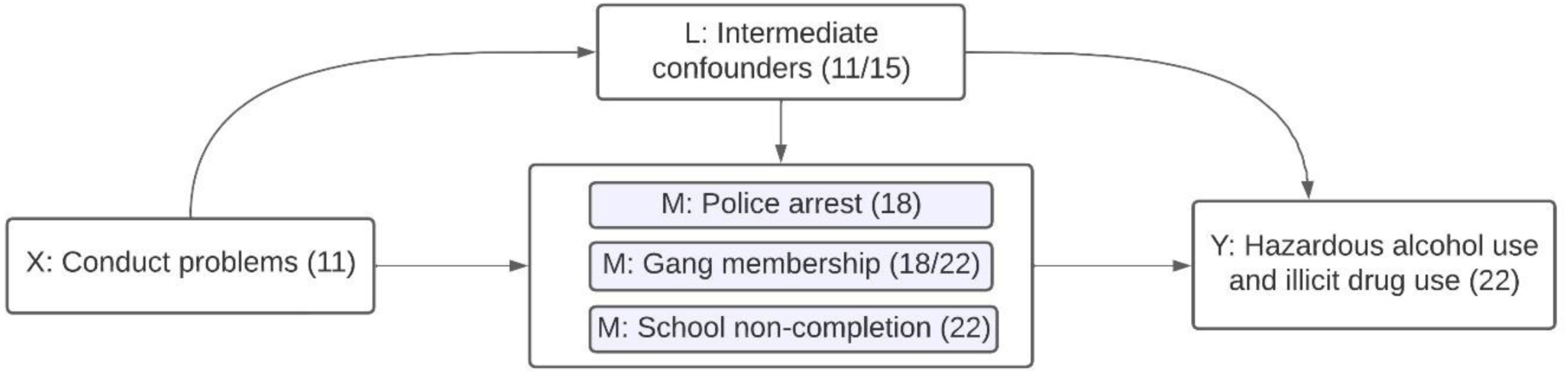
Directed Acyclic Graph showing hypothesised causal pathways between conduct problems and substance use (hazardous alcohol use and illicit drug use) through mediating “snares” (police arrest, gang membership, and school non-completion) Note: Baseline confounders (not shown on figure but assumed to confound all paths) included sex, sociodemographic risk factors, and health risk factors (assessed perinatally) and maternal depression, father-child relationship, mother-child relationship, parental alcohol consumption, parental smoking, parental separation, neighbourhood safety, and hyperactivity problems (assessed at age 11); intermediate confounders included peer deviance (assessed at age 11) and frequency of cigarette smoking, frequency of alcohol consumption, peer drug use, and emotional problems (assessed at age 15); hyperactivity problems were considered as baseline confounder in secondary analyses, and emotional problems were considered as an intermediate confounder in secondary analyses as these constructs represent comorbidity with conduct problems and the causal direction between them is less clear compared to other baseline/ intermediate confounders

#### Sensitivity analyses

First, we re-ran the mediation models making the assumption that police arrest (by age 18 years) and gang membership (at age 18 or 22 years) caused school non-completion (by age 22 years) i.e. we performed a single mediator model for school non-completion with police arrest and gang membership as additional intermediate confounders. Second, we re-ran all analyses using complete case rather than imputed data (further details provided below).

### Missing data

Of the 5,249 newborns included in the cohort, 45% (*N* = 2,342) had complete data on all analysis variables (see Figure S1, available online, for a flow chart of retention). Table S1, available online, provides a comparison of the complete case sample to the sample with missing information in at least one of the analysis variables (n = 2,907) on the exposure and baseline confounders. As shown in Table S1, high conduct problems at age 11 was associated with missingness, as was female sex, a higher sociodemographic and health risk score, higher maternal depression symptoms, no parental separation, no parental alcohol use, poorer father-child and mother-child relationship, and higher hyperactivity problems.

To increase sample size, we performed multivariate imputation by chained equations to impute missing data on all variables, across all assessments up to our starting sample (those that provided data on alcohol use and drug use from at least one time point between age 11 and age 22 years; *N* = 4,599). This starting sample was chosen to ensure we had strong auxiliary variables available to impute substance use at age 22 years. For a detailed description of multiple imputation, see Supplement 3, available online. Imputation and subsequent analysis code for the mediation models can be found on GitHub: https://github.com/gemmahammerton/gformula_1993_Pelotas. All analyses in the main text were performed on imputed data.

## Results

### Descriptive statistics

Of 4,599 children included in the imputed analyses, 51% were female, 77% had White mothers, and 23% had Black or mixed race mothers. Table 1 shows descriptive statistics for all analysis variables for the whole sample, using both imputed (*N* = 4,599) and complete case data (*N* = 2,342). Based on imputed data, 31% of children at age 11 years had high conduct problems, 4% of young people had been arrested or detained by the police by age 18 years, 2% had been members of a gang in the previous year at age 18 or 22 years, and 43% had not completed high school (passed all school grades) by age 22 years. 22% of young people reported hazardous alcohol consumption at age 22 years, and 14% reported current use of illicit drugs.

**Table 1.** Descriptive statistics for analysis variables for the whole sample using imputed and complete case data.

|  | Imputed data<br>(N = 4,599) | Complete case data<br>(N = 2,342) |
| --- | --- | --- |
| Variable | % / Mean (95% CI) | % / Mean (95% CI) |
| **Exposure (age 11)** |  |  |
| Conduct problems (high; 4+) | 31% (29% to 32%) | 27% (26% to 29%) |
| **Mediators (age 18-22)** |  |  |
| Police arrest (yes) | 4% (3% to 4%) | 3% (2% to 3%) |
| Gang membership (yes) | 2% (2% to 3%) | 1% (1% to 2%) |
| School non-completion (yes) | 43% (42% to 45%) | 36% (34% to 38%) |
| **Outcomes (age 22)** |  |  |
| Hazardous alcohol use | 22% (21% to 24%) | 21% (19% to 22%) |
| Illicit drug use | 14% (13% to 15%) | 13% (12% to 14%) |
| **Baseline confounders (perinatal)** |  |  |
| Female sex | 51% (49% to 52%) | 54% (52% to 56%) |
| Sociodemographic risk score (0 - 5) | 0.90 (0.87 to 0.93) | 0.78 (0.74 to 0.82) |
| Health risk score (0 - 6) | 1.56 (1.52 to 1.59) | 1.51 (1.47 to 1.56) |
| **Baseline confounders (age 11)** |  |  |
| Maternal depression (0 - 20) | 5.69 (5.56 to 5.82) | 5.47 (5.30 to 5.65) |
| Fear of the neighbourhood (yes) | 12% (11% to 13%) | 11% (10% to 13%) |
| Parental separation (yes) | 57% (56% to 59%) | 54% (52% to 56%) |
| Parental smoking (yes) | 73% (72% to 74%) | 72% (70% to 74%) |
| Parental alcohol use (yes) | 68% (66% to 69%) | 69% (67% to 71%) |
| Father-child relationship (1 (worst) - 5) | 4.10 (4.06 to 4.13) | 4.15 (4.10 to 4.20) |
| Mother-child relationship (1 (worst) - 5) | 4.43 (4.40 to 4.46) | 4.49 (4.46 to 4.53) |
| **Comorbidity (age 11)** |  |  |
| Hyperactivity problems (0 - 10) | 4.30 (4.21 to 4.39) | 4.10 (3.97 to 4.22) |
| **Intermediate confounders (age 11-15)** |  |  |
| Peer deviance (yes) | 47% (45% to 48%) | 45% (43% to 47%) |
| Frequency of cigarette smoking (1 to 5 days in last month) | 6% (5% to 7%) | 5% (4% to 5%) |
| Frequency of alcohol use (1 to 5 days in last month) | 25% (24% to 26%) | 24% (23% to 26%) |
| Peer drug use (yes) | 13% (12% to 14%) | 12% (11% to 14%) |
| **Comorbidity (age 15)** |  |  |
| Emotional problems (0 - 10) | 3.93 (3.84 to 4.01) | 3.82 (3.71 to 3.92) |

### Associations between conduct problems (exposure) and police arrest, gang membership and school non-completion (mediators)

Figure 2 shows associations between the exposure (high conduct problems) and the mediators (police arrest, gang membership, and school non-completion) before and after adjusting for confounders. After adjusting for baseline confounders and hyperactivity problems, those with high childhood conduct problems had slightly higher odds of being arrested by the police by age 18 [OR (95% CI) = 1.45 (0.97, 2.16)] and not completing school by age 22 [OR (95% CI) = 1.46 (1.22, 1.74)] compared to those with low childhood conduct problems. There was no evidence of an association between conduct problems and gang membership after adjusting for baseline confounders and hyperactivity problems [OR (95% CI) = 1.17 (0.70, 1.95)].

**Figure 2.**
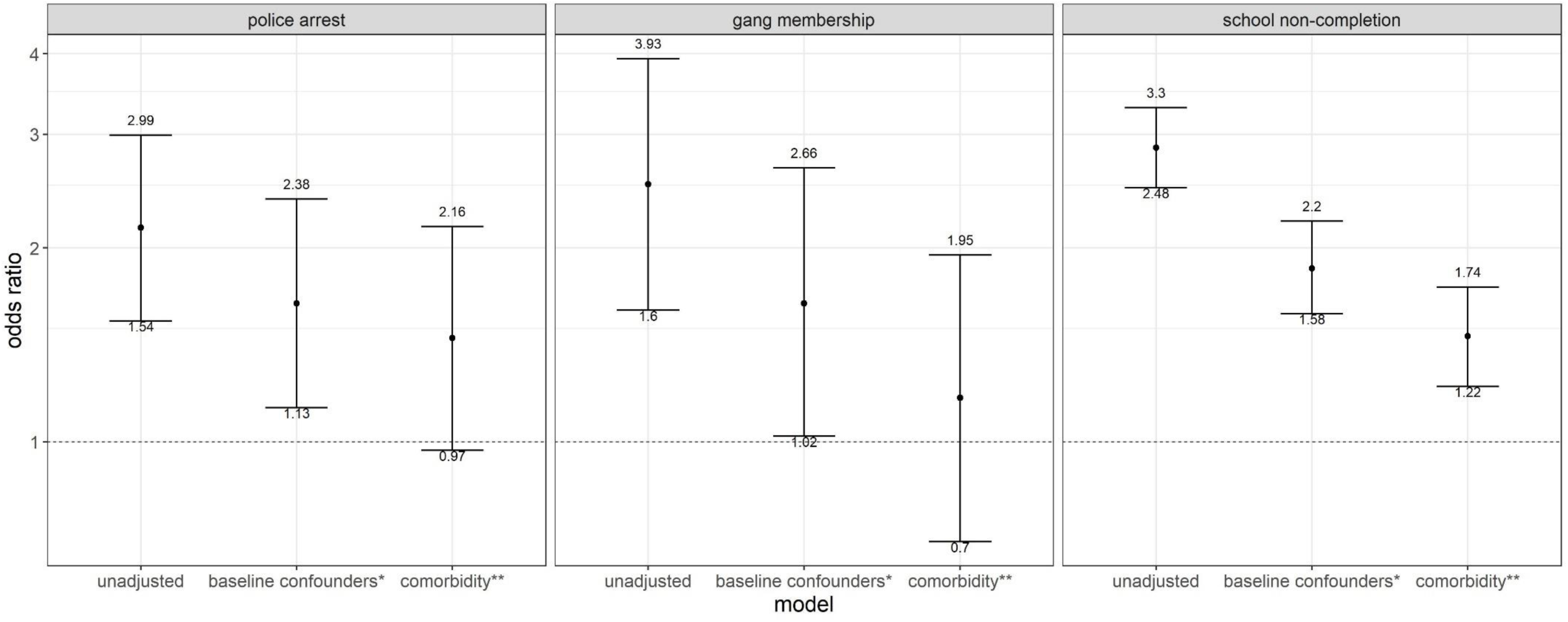
Associations between the exposure (conduct problems at age 11 years) and the mediators (police arrest, gang membership, and school non-completion between age 18 and 22 years) using imputed data; *N* = 4,599 Figure showing odds ratio and 95% confidence interval; * adjusted for baseline confounders including sex, score of sociodemographic risk factors, score of health risk factors (all measured perinatally), maternal depression, father/child relationship, mother/child relationship, parental alcohol consumption, parental smoking, parental separation, neighbourhood safety (all measured at age 11); **adjusted for baseline confounders listed above, and additionally adjusted for hyperactivity problems at age 11

### Associations between police arrest, gang membership and school non-completion (mediators) and hazardous alcohol use and illicit drug use (outcomes)

Univariable associations for baseline confounders with mediators and outcomes are shown in Table S3, available online, and univariable associations for intermediate confounders are shown in Table S4.

Table 2 shows associations between the mediators and the outcomes before and after adjusting for confounders (including hyperactivity and emotional problems). After adjusting for all confounders, those who had been arrested [OR (95% CI) = 1.60 (1.08, 2.38)] and those in a gang [OR (95% CI) = 1.88 (1.07, 3.30)] had slightly higher odds of hazardous alcohol use at age 22. There was no evidence of an association between school non-completion and hazardous alcohol use after adjusting for confounders [OR (95% CI) = 1.10 (0.90, 1.33)].

**Table 2.**
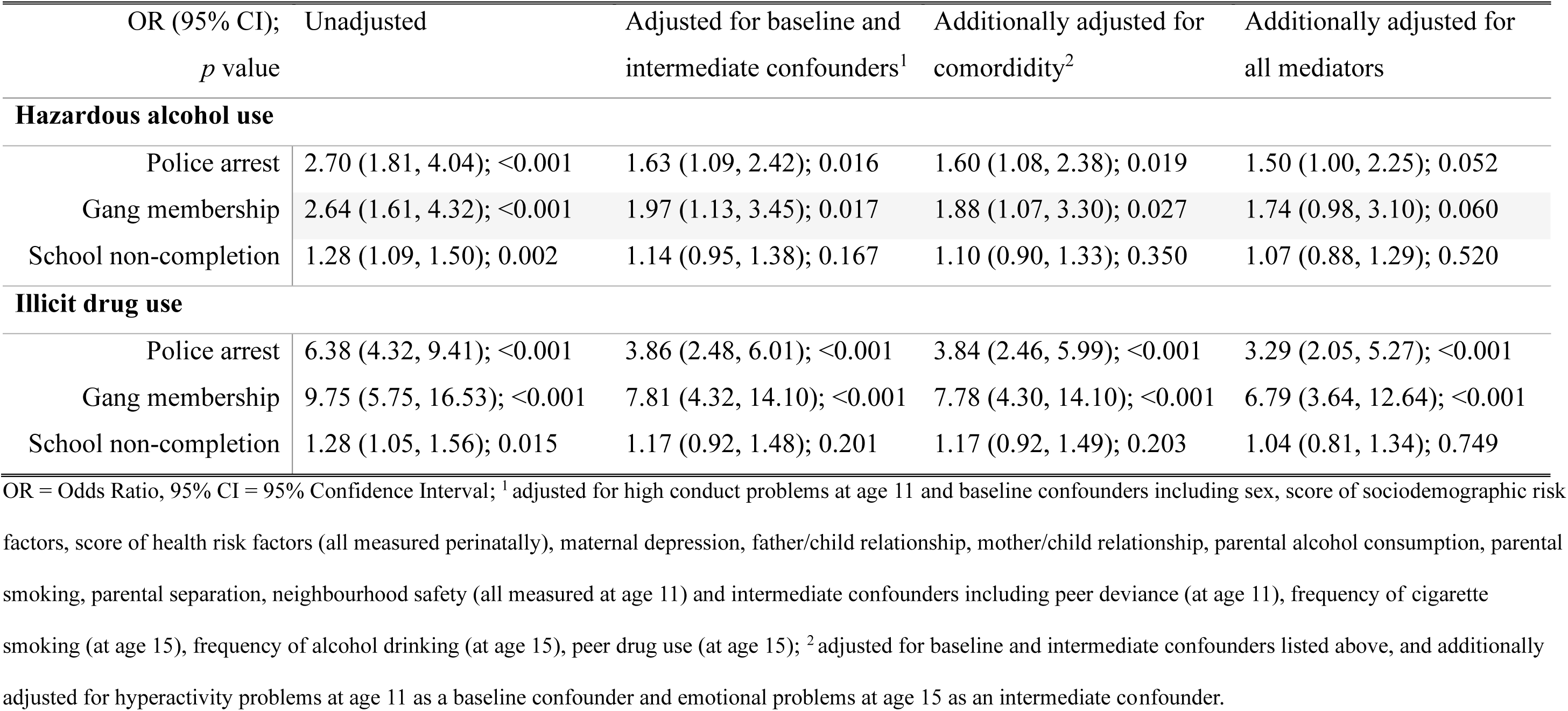
Associations between the mediators (police arrest, gang membership, and school non-completion between age 18 and 22 years) and the outcomes (hazardous alcohol use and illicit drug use at age 22 years) using imputed data; *N* = 4,599.

| OR (95% CI);<br>$p$ value | Unadjusted | Adjusted for baseline and<br>intermediate confounders<sup>1</sup> | Additionally adjusted for<br>comorbidity<sup>2</sup> | Additionally adjusted for<br>all mediators |
| --- | --- | --- | --- | --- |
| **Hazardous alcohol use** |  |  |  |  |
| Police arrest | 2.70 (1.81, 4.04); <0.001 | 1.63 (1.09, 2.42); 0.016 | 1.60 (1.08, 2.38); 0.019 | 1.50 (1.00, 2.25); 0.052 |
| Gang membership | 2.64 (1.61, 4.32); <0.001 | 1.97 (1.13, 3.45); 0.017 | 1.88 (1.07, 3.30); 0.027 | 1.74 (0.98, 3.10); 0.060 |
| School non-completion | 1.28 (1.09, 1.50); 0.002 | 1.14 (0.95, 1.38); 0.167 | 1.10 (0.90, 1.33); 0.350 | 1.07 (0.88, 1.29); 0.520 |
| **Illicit drug use** |  |  |  |  |
| Police arrest | 6.38 (4.32, 9.41); <0.001 | 3.86 (2.48, 6.01); <0.001 | 3.84 (2.46, 5.99); <0.001 | 3.29 (2.05, 5.27); <0.001 |
| Gang membership | 9.75 (5.75, 16.53); <0.001 | 7.81 (4.32, 14.10); <0.001 | 7.78 (4.30, 14.10); <0.001 | 6.79 (3.64, 12.64); <0.001 |
| School non-completion | 1.28 (1.05, 1.56); 0.015 | 1.17 (0.92, 1.48); 0.201 | 1.17 (0.92, 1.49); 0.203 | 1.04 (0.81, 1.34); 0.749 |

OR = Odds Ratio, 95% CI = 95% Confidence Interval; <sup>1</sup> adjusted for high conduct problems at age 11 and baseline confounders including sex, score of sociodemographic risk factors, score of health risk factors (all measured perinatally), maternal depression, father/child relationship, mother/child relationship, parental alcohol consumption, parental smoking, parental separation, neighbourhood safety (all measured at age 11) and intermediate confounders including peer deviance (at age 11), frequency of cigarette smoking (at age 15), frequency of alcohol drinking (at age 15), peer drug use (at age 15); <sup>2</sup> adjusted for baseline and intermediate confounders listed above, and additionally adjusted for hyperactivity problems at age 11 as a baseline confounder and emotional problems at age 15 as an intermediate confounder.

After adjusting for confounders, those who had been arrested [OR (95% CI) = 3.84 (2.46, 5.99)] and those in a gang [OR (95% CI) = 7.78 (4.30, 14.10)] had higher odds of illicit drug use at age 22. There was no evidence of an association between school non-completion and illicit drug use after adjusting for confounders [OR (95% CI) = 1.17 (0.92, 1.49)].

### Indirect effects of conduct problems (exposure) on hazardous alcohol use and illicit drug use (outcomes) via police arrest, gang membership and school non-completion (mediators)

Table 3 shows the total causal effect, natural direct effect and natural indirect effect of conduct problems on hazardous alcohol use and illicit drug use via all three mediators simultaneously. After adjusting for baseline and intermediate confounders, there was weak evidence for a small total effect of conduct problems on hazardous alcohol use [OR (95% CI) = 1.18 (1.00, 1.39)] but little evidence for a total effect of conduct problems on illicit drug use [OR (95% CI) = 1.05 (0.86, 1.30)]. There was weak evidence for a small natural indirect effect of conduct problems on hazardous alcohol use [OR (95% CI) = 1.03 (1.00, 1.06)] and illicit drug use [OR (95% CI) = 1.08 (1.01, 1.17)] via all three mediators.

**Table 3.**
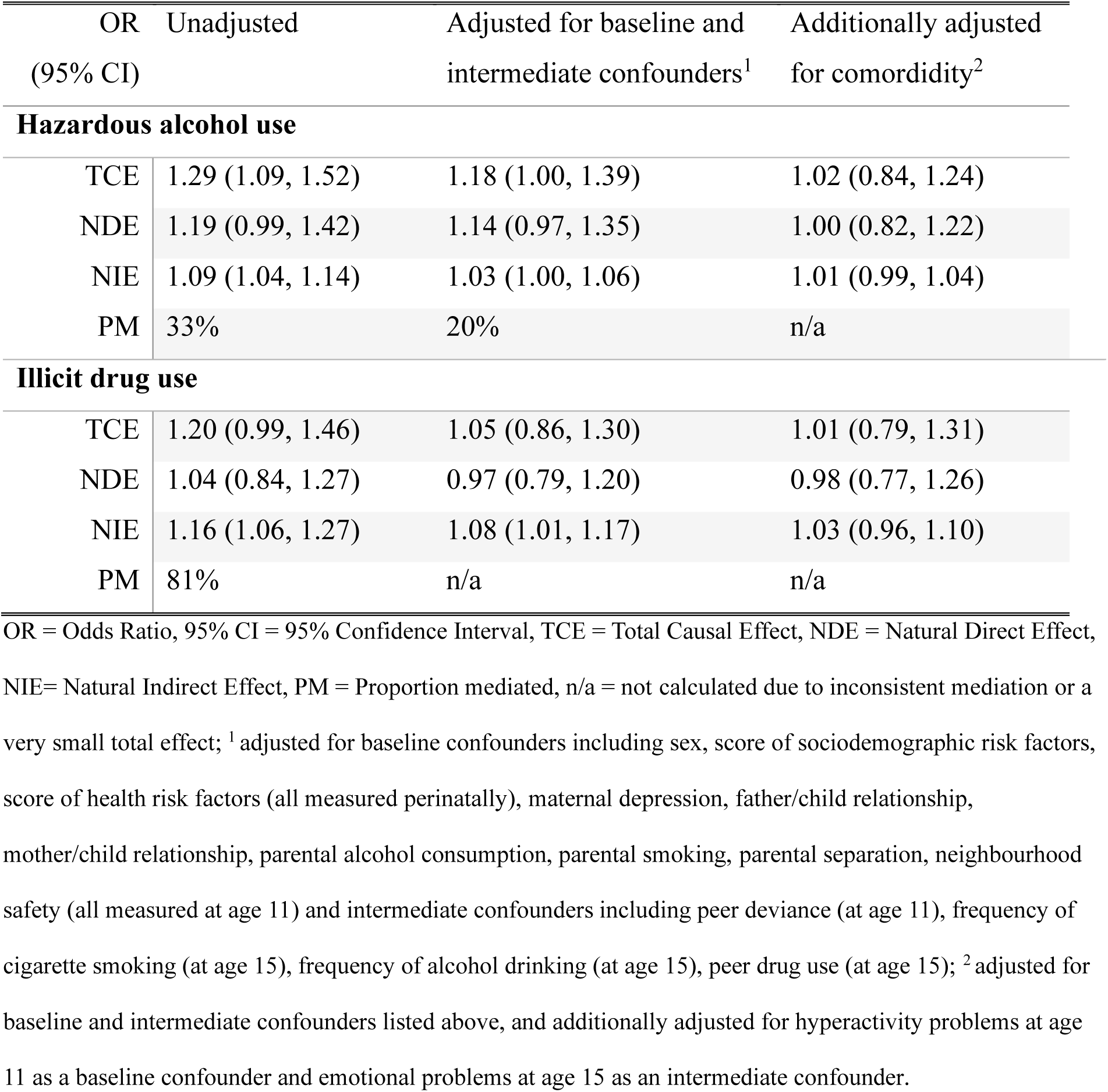
Mediation models including all mediators (police arrest, gang membership, and school non-completion) simultaneously using imputed data; *N* = 4,599.

| OR<br>(95% CI) | Unadjusted | Adjusted for baseline and<br>intermediate confounders<sup>1</sup> | Additionally adjusted<br>for comorbidity<sup>2</sup> |
| --- | --- | --- | --- |
| **Hazardous alcohol use** |  |  |  |
| TCE | 1.29 (1.09, 1.52) | 1.18 (1.00, 1.39) | 1.02 (0.84, 1.24) |
| NDE | 1.19 (0.99, 1.42) | 1.14 (0.97, 1.35) | 1.00 (0.82, 1.22) |
| NIE | 1.09 (1.04, 1.14) | 1.03 (1.00, 1.06) | 1.01 (0.99, 1.04) |
| PM | 33% | 20% | n/a |
| **Illicit drug use** |  |  |  |
| TCE | 1.20 (0.99, 1.46) | 1.05 (0.86, 1.30) | 1.01 (0.79, 1.31) |
| NDE | 1.04 (0.84, 1.27) | 0.97 (0.79, 1.20) | 0.98 (0.77, 1.26) |
| NIE | 1.16 (1.06, 1.27) | 1.08 (1.01, 1.17) | 1.03 (0.96, 1.10) |
| PM | 81% | n/a | n/a |

OR = Odds Ratio, 95% CI = 95% Confidence Interval, TCE = Total Causal Effect, NDE = Natural Direct Effect, NIE = Natural Indirect Effect, PM = Proportion mediated, n/a = not calculated due to inconsistent mediation or a very small total effect; <sup>1</sup> adjusted for baseline confounders including sex, score of sociodemographic risk factors, score of health risk factors (all measured perinatally), maternal depression, father/child relationship, mother/child relationship, parental alcohol consumption, parental smoking, parental separation, neighbourhood safety (all measured at age 11) and intermediate confounders including peer deviance (at age 11), frequency of cigarette smoking (at age 15), frequency of alcohol drinking (at age 15), peer drug use (at age 15); <sup>2</sup> adjusted for baseline and intermediate confounders listed above, and additionally adjusted for hyperactivity problems at age 11 as a baseline confounder and emotional problems at age 15 as an intermediate confounder.

After additionally adjusting for hyperactivity (at age 11) as a baseline confounder and emotional problems (at age 15) as an intermediate confounder, all effects weakened. There was no longer evidence for a total effect of conduct problems on hazardous alcohol use [OR (95% CI) = 1.02 (0.84, 1.24)], or a natural indirect effect of conduct problems on hazardous alcohol use [OR (95% CI) = 1.01 (0.99, 1.04)] or illicit drug use [OR (95% CI) = 1.03 (0.96, 1.10)] via all three mediators.

### Sensitivity analyses

Table S5, available online, shows the total causal effect, natural direct effect and natural indirect effect of conduct problems on hazardous alcohol use and illicit drug use via school non-completion, treating police arrest and gang membership as intermediate confounders. There was no evidence for an indirect effect via school non-completion, for either hazardous alcohol use or illicit drug use after adjusting for confounders.

Tables S6 to S8, available online, show results for all analyses using complete case data (*N* = 2,342). In general, odds ratios were slightly stronger but confidence intervals were wider using the complete case sample; however, conclusions were unchanged.

## Discussion

### Summary of findings

Using a large, population-based birth cohort in Brazil, we found that high childhood conduct problems were weakly associated with being arrested by the police by age 18 years and strongly associated with not completing school by age 22 years after adjusting for potential confounders, including childhood hyperactivity problems. We also found an association between childhood conduct problems and gang membership in late adolescence, but this weakened substantially after accounting for childhood hyperactivity problems. Being arrested by the police and gang membership were strongly associated with illicit drug use and weakly associated with hazardous alcohol use at age 22 years, after adjusting for confounders (including childhood conduct, hyperactivity, and emotional problems). However, there was no association between school non-completion and either hazardous alcohol use or illicit drug use after adjusting for confounders. Results from the mediation analysis showed weak evidence for a total effect of conduct problems on hazardous alcohol use but little evidence for a total effect of conduct problems on illicit drug use after adjusting for confounders. We found weak evidence for a natural indirect effect of conduct problems on hazardous alcohol use and illicit drug use via all three mediators; however, after additionally adjusting for hyperactivity and emotional problems, there was no longer evidence of an indirect effect on either outcome.

### Strengths and limitations

Due to the longitudinal design and wealth of data in the 1993 Pelotas Birth Cohort, we were able to examine mediators of the association between conduct problems at age 11 and substance use over 10 years later, whilst adjusting for potential baseline and intermediate confounders across multiple domains, including sociodemographic, individual, peer, family, and the community. However, our results need to be interpreted in light of several limitations. First, in our population-based sample, few people joined a gang or were arrested by the police (2% and 4%, respectively); therefore, the weak evidence for associations could be explained by a lack of power and other factors more common among children with conduct problems might be more important mediators for the relationship between conduct problems and substance use.

Second, even though we adjusted for a wide range of confounders, there might be some residual confounding present in our analysis due to either unmeasured confounders (such as genetic risk) or confounders measured with error. Third, as with most cohort studies, there was selective attrition over time, with missingness associated with conduct and hyperactivity problems, alongside sociodemographic and family-based confounders.

However, we used multiple imputation to address missing data incorporating a wide range of strong auxiliary variables to make the missing-at-random assumption plausible and conclusions using complete case and imputed data were similar.

Fourth, although the exposure (conduct problems) was assessed using parent-report, the mediators and outcomes were assessed using self-report meaning that results could be subject to reporting bias and shared-rater bias. Additionally, although the exposure and outcomes were assessed using established scales that have been validated in Brazil, each mediator was assessed using a single item. Finally, the measure of school non-completion captures both dropping out of school and grade repetition. It may be that school dropout is a more important risk factor for substance use than grade repetition, but we were not able to distinguish between these related constructs.

### Comparison with previous literature

We found that childhood conduct problems were strongly associated with not completing school by age 22 years after adjusting for confounders, including childhood hyperactivity problems. This finding partially supports a previous study using a school-based community cohort of Brazilian children and adolescents (aged 6 to 14 years) which found that externalising disorders (including attention deficit and hyperactivity, conduct and oppositional-defiant disorders) were associated with grade repetition and age-grade distortion three years later^15^, although not with school dropout. This finding also supports evidence from HIC showing an association between childhood conduct problems and school non-completion^16–20^. Contrary to the conclusion in a recent systematic review^19^, we found evidence of an association between conduct problems and school non-completion, even after adjusting for hyperactivity problems. However, although high childhood conduct problems were associated with both police arrest and gang membership after adjusting for confounders, these associations weakened after adjusting for childhood hyperactivity problems, particularly for gang membership. This supports previous reviews of the literature which highlight the importance of risk-taking, impulsivity, and low self-control as risk factors for joining a gang^9,24,45^. However, any one risk factor in isolation is unlikely to be sufficient to identify young people at high or low risk of joining a gang; alternatively, the total accumulation of risk factors or exposure to risk factors across multiple domains is important^24,46,47^.

In the current study, there was no association between school non-completion and either hazardous alcohol use or illicit drug use after adjusting for confounders. This is in contrast to a study of American adolescents which found an association between school failure and alcohol use one year later^21^. The lack of association in the current study could be explained by the wide range of confounders adjusted for (given that there was evidence for an association in the unadjusted analyses). Alternatively, it could be due to the age of the young people, given that those who have completed school may be attending university at age 22, where levels of substance use are generally high. Despite the lack of association between school non-completion and substance use, it is still likely that school non-completion is an important snare in Brazil, that may trap those with childhood conduct problems into involvement with violent and non-violent crime in early adulthood^23^.

In contrast to the findings for school non-completion, we found that being arrested by the police and gang membership were weakly associated with hazardous alcohol use, and strongly associated with illicit drug use at age 22 years, even after adjusting for confounders (including childhood conduct, hyperactivity, and emotional problems). This supports results from previous studies that found associations between gang membership^9,25,48,49^ and police arrest^28^ with problematic substance use.

### Conclusions and future directions

The findings from the current study highlight the importance of school professionals being aware of the risk for school non-completion for those with childhood conduct problems, and collaboration between education and health sectors in Brazil. Early intervention to support those with childhood conduct problems to continue with their education is a priority. However, it is also important to consider that many behavioral problems have to do with not adapting to the school system, which is not a problem in itself for the child or adolescent, but our inability to think about education in other ways. Joining gangs can be a way of protesting, finding peers who may also not agree with the ideas of others, of the institutions.

The findings also highlight the importance of early intervention and treatment for hyperactivity (in addition to conduct problems) to prevent long-term negative outcomes such as gang membership, criminal activity, and substance use in Brazil. Given the high rates of comorbidity of conduct and hyperactivity problems, it is difficult to examine the independent effects and preventing or treating both is a priority^17,29^. Finally, given the strong associations observed between gang membership and police arrest with substance use (particularly illicit drug use), programmes and policies designed to reduce substance use in Brazil should focus on young people involved in gangs, and in the criminal justice system.

In the current study, we found little evidence for an indirect effect of childhood conduct problems on substance use in early adulthood via school non-completion, gang membership, and police arrest after accounting potential confounders including comorbidity. The weak evidence for mediating effects might be because the causes of problematic substance use in children with conduct problems operate early in life^6^. Alternatively, there may be other mediators, such exposure to trauma or the influence of deviant peers, which might have a stronger influence on later substance use^50^. Future research is needed to examine mediators of the associations between childhood hyperactivity problems and later substance use in Brazil, and to examine the potential snares that may trap children with conduct problems into other negative long-term outcomes such as criminal behavior and unemployment.

## Data Availability

Access to data from the 1993 Pelotas Birth Cohort is through a system of managed open access (https://epidemio-ufpel.org.br/cohort-1993/).

https://github.com/gemmahammerton/gformula_1993_Pelotas

## Funding

Dr. Hammerton is supported by the National Institute for Health Research (NIHR) Health Protection Research Unit in Behavioural Science and Evaluation (HPRU BSE) at the University of Bristol, in partnership with the UK Health Security Agency (UKHSA). This research was also funded by a Sir Henry Wellcome Postdoctoral Fellowship (209138/Z/17/Z) awarded to Dr. Hammerton. Drs. Hammerton and Heron are members of the MRC Integrative Epidemiology Unit at the University of Bristol (MC_UU_00011/7). This research was funded in whole, or in part, by the Wellcome Trust (209138/Z/17/Z; 210735_A_18_Z). For the purpose of Open Access, the authors have applied a CC BY public copyright licence to any Author Accepted Manuscript version arising from this submission. Professor Murray is supported by a Wellcome Trust Investigator Award (210735_A_18_Z). Professors Menezes, Gonçalves, Wehrmeister, and Murray are supported by the Brazilian National Research Council (CNPq).

Drs. Heron and Hammerton, PhD, served as the statistical experts for this research.

## Acknowledgements

This article is based on data from the study "Pelotas Birth Cohort, 1993" conducted by Postgraduate Program in Epidemiology at Universidade Federal de Pelotas with the collaboration of the Brazilian Public Health Association (ABRASCO). From 2004 to 2013, the Wellcome Trust supported the 1993 birth cohort study. The European Union, National Support Program for Centers of Excellence (PRONEX), the Brazilian National Research Council (CNPq), and the Brazilian Ministry of Health supported previous phases of the study. The 22-year follow-up was supported by the Science and Technology Department / Brazilian Ministry of Health, with resources transferred through the Brazilian National Council for Scientific and Technological Development (CNPq), grant 400943/2013-1.

Dr. Hammerton and Professor Hickman would like to acknowledge support from the National Institute for Health Research (NIHR) Health Protection Research Unit in Behavioural Science and Evaluation (HPRU BSE) at the University of Bristol. The views expressed are those of the authors and not necessarily those of the NIHR, the Department of Health and Social Care or UK Health Security Agency (UKHSA).

All authors have reported no biomedical financial interests or potential conflicts of interest.

## Supplement 1. Details on the baseline and intermediate confounders

### Baseline and intermediate confounders

Baseline confounders included sex, a score of sociodemographic and health risk factors for conduct problems, maternal depression, the quality of the father-child and mother-child relationship, parental alcohol consumption, parental smoking status, parental separation, and neighbourhood safety. These were chosen based on evidence from previous literature of associations with conduct problems ^1–4^, the potential snares ^2,4–8^, and substance use ^9–13^.

Sociodemographic and health risk factors were assessed by maternal report during the perinatal period. Sociodemographic factors included maternal age (< 20 years/≥20 years), low maternal education (yes/no; referring to 0–8 vs≥ 9 years of schooling), marital status (single mother/ with partner), three or more siblings (yes/no) and family income (lowest quintile/ second-fifth quintiles). The cumulative number of sociodemographic risk factors was summed, up to five, for each child as has been done previously ^2^, with five indicating the highest risk for conduct problems and substance use. Health risk factors included unplanned pregnancy (yes/no), mother ever smoked in pregnancy (yes/no), mother used alcohol in pregnancy (yes/no), maternal urinary infection during pregnancy (yes/no), intrauterine growth restriction (yes/no; referring to < 10th percentile/≥ 10th percentile for gestational age and sex, according to the reference curve developed by Kramer and colleagues (Kramer et al, 2001) and premature birth < 37 weeks (yes/no). Again, the cumulative number of health risk factors was summed, up to six, for each child as has been done previously ^2^.

All other baseline confounders were assessed when the child was 11 years old. Maternal depression was assessed with mothers using the Self Report Questionnaire, which has been validated in Brazil ^14^. The quality of the mother-child and father-child relationship was assessed with child-report questions including the following response options: bad, regular, good, very good, excellent. Parental alcohol consumption was assessed using two maternal-report questions, capturing whether she and/or her husband never, not currently or currently drink alcohol. Likewise, parental smoking was assessed using two maternal-report questions, one addressing the mother’s lifetime smoking status and one addressing the father’s lifetime smoking status (ever versus never). Parental separation was assessed using a maternal-report question about the child’s biological father living in the same household, and a child-report question about parental divorce. A binary variable was created, were either child-report or maternal-report of the father not living in the same household were classed as parental separation. Neighbourhood safety was assessed using a maternal-report question asking whether she was afraid of living in the neighbourhood (yes/no).

Intermediate confounders included peer deviance, frequency of cigarette smoking and alcohol consumption, and peer drug use. Intermediate confounders were chosen based on evidence that they confound the associations between the potential snares and substance use ^5–8,10,12,15,16^, and are also causally downstream from childhood conduct problems (our exposure) ^5,12,17–19^.

Peer deviance was assessed at age 11 by asking the young person’s mother if she considered her child’s friends as having a bad influence or being bad company (yes/no). Frequency of cigarette smoking and frequency of alcohol consumption were assessed at age 15 by child-report. The questionnaire item for smoking had five response categories: ‘Never smoked cigarettes, not smoked in the last month, 1 to 5 days, 6 to 9 days, ten or more days and every day in the last month’ and the questionnaire item for alcohol had five response categories: ‘Never drank alcohol, not drank alcohol in the last month, one to five days, six to nine days, ten or more days and every day in last month’. For both items, the last three response categories were combined to the category “current smoking” or “current drinking”, respectively, due to small cell counts. Peer drug use was assessed by child-report at age 15. A binary variable was created indicating if friends or anyone in the same class as the young person used cannabis, glue, solvents, or cocaine.

### Comorbidity: hyperactivity problems (age 11) and emotional problems (age 15)

In secondary analyses, hyperactivity was considered as an additional baseline confounder based on evidence for associations with conduct problems ^1,3,4,20^, the potential snares ^6,7,19,20^, and substance use ^19,21–23^. Hyperactivity problems were assessed by maternal-report at child age 11 years using the five-item hyperactivity problems subscale from the Strengths and Difficulties Questionnaire (SDQ) ^24^. The five items in the questionnaire include ‘restless, overactive, cannot stay still for long’, ‘constantly fidgeting or squirming’, ‘easily distracted, concentration wanders’, ‘thinks things out before acting’ and ‘sees tasks through to the end, good attention span’. Responses for each item are ‘Not true’, ‘Somewhat true’ and ‘Certainly true’. The items ‘thinks things out’ and ‘sees tasks through’ were reverse coded. All items were summed together (range 0 to 10).

In secondary analyses, emotional problems were considered as an additional intermediate confounder given that they may be a precursor for the potential snares and substance use ^23,25–28^, and are also causally downstream from childhood conduct problems (our exposure) ^19,29^. Emotional problems were assessed by maternal-report at age 15 years using the five-item emotional problems subscale from the SDQ. The five items in the questionnaire include ‘often complains of headaches, stomach-aches or sickness’, ‘many worries, often seems worried’, ‘often unhappy, down-hearted or tearful’, ‘nervous or clingy in new situations, easily loses confidence’ and ‘many fears, easily scared’. Responses for each item are ‘Not true’, ‘Somewhat true’ and ‘Certainly true’. All items were summed together (range 0 to 10).

## Supplement 2. Further details on the statistical analyses including i) logistic regression models and ii) the counterfactual approach to mediation

### Logistic regression models

We conducted unadjusted and adjusted logistic regressions in stages to examine exposure-mediator and mediator-outcome associations. For exposure-mediator regressions, we first adjusted for baseline confounders, followed by baseline confounders including hyperactivity problems. For mediator-outcome regressions, we first adjusted for baseline and intermediate confounders, followed by these confounders including hyperactivity and emotional problems, followed by a regression model simultaneously including all three mediators, and all confounders. All regression models were governed by the same Directed Acyclic Graph, hence for mediator-outcome regressions, the exposure (conduct problems) becomes another confounder. Caution is required when directly comparing across these logistic regression models, each with different sets of confounders, given that the non-collapsibility of the odds ratio with a common binary outcome (22% for hazardous alcohol use and 14% for illicit drug use) means that the difference between an unadjusted and adjusted odds ratio can be due to both confounder adjustment and non-collapsibility ^30^.

### Counterfactual mediation

The counterfactual approach is based on conceptualising ‘potential outcomes’ for each individual [Y(*x*)] that would have been observed if particular conditions were met (i.e. had the exposure X been set to the value *x* through some intervention) – regardless of the conditions that were in fact met for each individual ^31^. In mediation analyses, four assumptions are made with respect to confounding. These include no unmeasured confounders for any of the paths (exposure-outcome, exposure-mediator and mediator-outcome) and no measured or unmeasured confounders for the association between mediator and outcome which lie on the causal pathway from the exposure. In the current study, we assume that the same set of baseline confounders (sex, a score of sociodemographic and health risk factors, maternal depression, the quality of the father-child and mother-child relationship, parental alcohol consumption, and smoking status, parental separation, neighbourhood safety, and hyperactivity problems) confound all paths, and these were assessed before or during the same assessment as the exposure.

The counterfactual approach to mediation using the parametric g-computation formula was performed to relax the assumption of no measured intermediate confounding and to include binary mediators and outcomes. This counterfactual approach simulates the mediator, outcome and intermediate confounders under each hypothetical intervention or “counter to the fact” scenario. Again, mediation models were performed in stages. First we performed unadjusted models. Next, we adjusted for baseline and intermediate confounders, and finally, we adjusted for all confounders including hyperactivity and emotional problems. Unlike the logistic regression models, -gformula-presents mediation effects as marginal odds ratios meaning that non-collapsibility of the odds ratio is no longer a consideration. We did not hypothesise an exposure-mediator interaction and therefore did not include it in our models. The proportion of the total causal effect that was mediated was calculated by dividing the log odds ratio for the natural indirect effect by the log odds ratio for the total causal effect, and multiplying by 100. We used a Monte Carlo sample size of 10,000 to minimise fluctuations in effect estimates, and a seed of 79.

The total causal effect (TCE) is the value the outcome would take if everybody had been exposed to high conduct problems versus everyone having low conduct problems. The natural direct effect (NDE) is the direct (unmediated) effect of the exposure on the outcome when the mediator takes the value it would take in the absence of the exposure. It is thus modelled as the direct effect of exposure X = 1 (high conduct problems) versus exposure X = 0 (low conduct problems) on outcome Y (e.g., hazardous alcohol use) if mediator M (e.g., gang membership) were set to whatever it would be for X = 0.

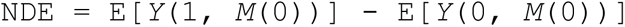

The NIE captures the effect of the exposure on the outcome that operates by changing the mediator. It is thus modelled as the effect on outcome Y (e.g., hazardous alcohol use) if the exposure were fixed at X = 1 and mediator M (e.g., gang membership) were changed from the level it would take if X = 0 to the level it would take if X = 1.

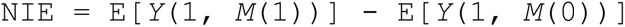

**Figure S1.**
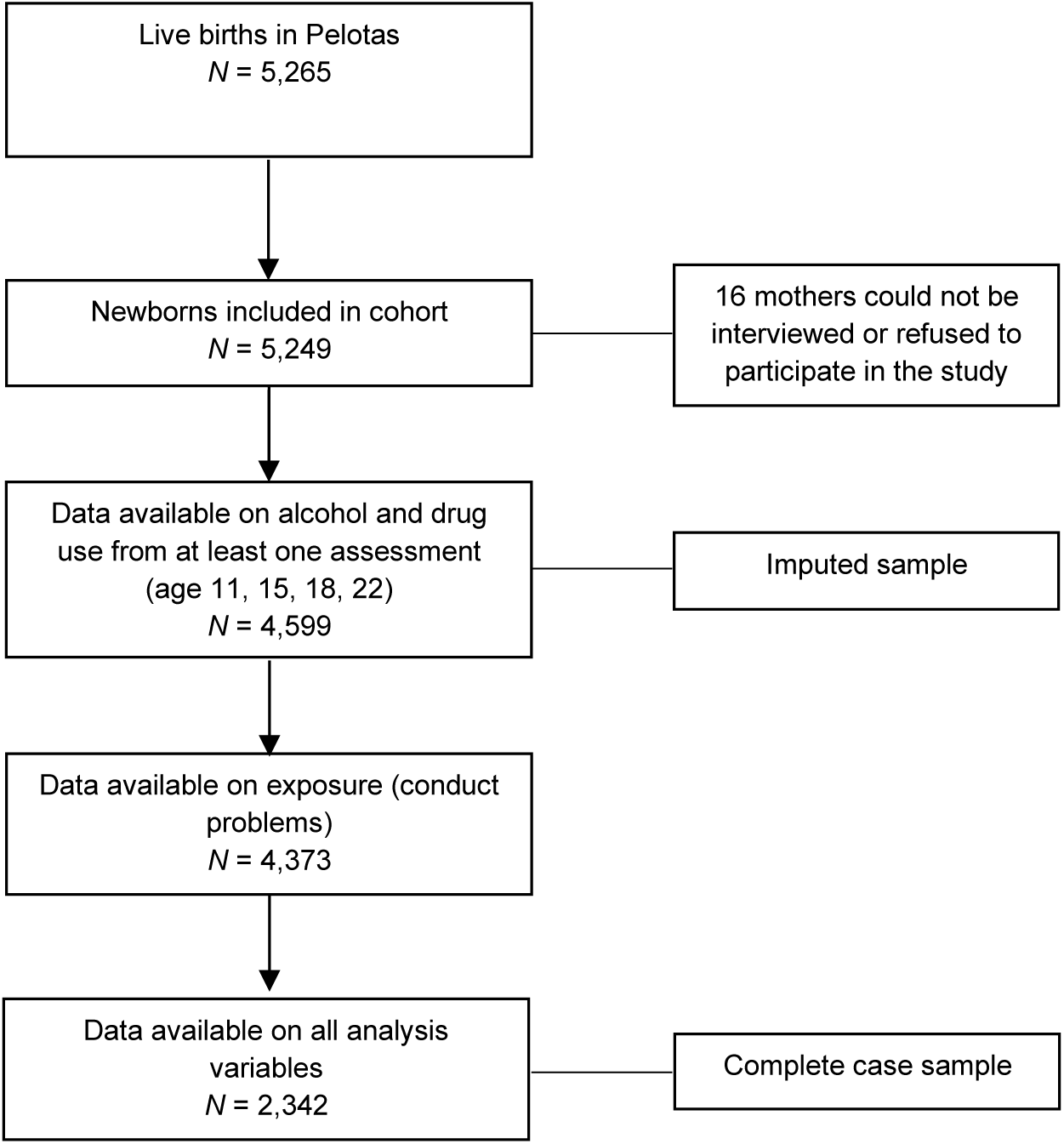
Flow chart of retention in the 1993 Pelotas Birth Cohort

**Table S1.** Comparison of complete cases (n = 2,342) and those with missing information in at least one of the analysis variables (n = 2,907)

|  | Proportion<br>missingness | Complete cases<br>*n* = 2342 |  | Cases with missing data<br>*n* = 2907 |  | Univariable association<br>with missingness |  |  |
| --- | --- | --- | --- | --- | --- | --- | --- | --- |
| Variable | % | *n* | (%) or<br>Mean (SD) | *n* | (%) or<br>Mean (SD) | OR | (95% CI) | p-value |
| **Exposure (age 11)** |  |  |  |  |  |  |  |  |
| Conduct problems (high; 4+) | 16% | 640. | (27.33) | 733. | (35.53) | 1.47 | (1.30 - 1.67) | <0.0001 |
| **Baseline confounders (perinatal)** |  |  |  |  |  |  |  |  |
| Female sex | <1% | 1268. | (54.14) | 1377. | (47.38) | 0.76 | (0.68 - 0.85) | <0.0001 |
| Sociodemographic risk score (0 - 5) | 0% | 2342 | 0.78 (0.92) | 2907 | 1.03 (1.04) | 1.30 | (1.23 - 1.37) | <0.0001 |
| Health risk score (0 - 6) | 11% | 2342 | 1.51 (1.07) | 2330 | 1.59 (1.07) | 1.07 | (1.02 - 1.13) | 0.009 |
| **Baseline confounders (at age 11)** |  |  |  |  |  |  |  |  |
| Maternal depression (0 - 20) | 16% | 2342 | 5.47 (4.36) | 2060 | 6.01 (4.70) | 1.03 | (1.01 - 1.04) | <0.0001 |
| Fear of the neighbourhood (yes) | 16% | 267. | (11.40) | 273. | (13.14) | 1.18 | (0.98 - 1.41) | 0.079 |
| Parental separation (yes) | 0% | 1271. | (54.27) | 1380. | (47.47) | 0.76 | (0.68 - 0.85) | <0.0001 |
| Parental smoking (yes) | 16% | 1687. | (72.03) | 1532. | (73.55) | 1.08 | (0.95 - 1.23) | 0.259 |
| Parental alcohol use (yes) | 16% | 1618. | (69.09) | 1364. | (65.48) | 0.85 | (0.75 - 0.96) | 0.011 |
| Father-child relationship (1 (worst) - 5) | 18% | 2342 | 4.15 (1.16) | 1937 | 4.03 (1.22) | 0.92 | (0.87 - 0.97) | 0.001 |
| Mother-child relationship (1 (worst) - 5) | 17% | 2342 | 4.49 (0.85) | 1997 | 4.35 (0.97) | 0.84 | (0.79 - 0.90) | <0.0001 |
| Hyperactivity problems (0 - 10) | 16% | 2342 | 4.10 (3.09) | 2066 | 4.58 (3.08) | 1.05 | (1.03 - 1.07) | <0.0001 |

N = number/frequency, OR = Odds Ratio, 95%CI = 95% Confidence Interval, SD = Standard deviation, complete = no missing information

## Supplement 3. Details on multiple imputation

To address missing data, we used multivariate imputation by chained equations^32^ to impute all incomplete analysis variables up to our starting sample (those that provided data on alcohol use and drug use from at least one time point between age 11 and age 22 years; N = 4,599). Despite the univariable association observed between conduct problems and missingness (suggesting that the exposure could be missing-not-at-random), we decided to impute the small amount of missing data on conduct problems within our starting sample (<5% missing) given the strong auxiliary data and wide range of confounders available for our imputation model, making the missing-at-random assumption plausible. First, we investigated the patterns of missing data, the percentage of missingness for each analysis variable and potential auxiliary variables and determined the distribution of missing variables (see Supplementary Table 2). Next, we performed multivariable regression analyses with each incomplete analysis variable as the outcome and potential auxiliary variables (such as earlier assessments of the mediators and outcomes) as the exposures to determine which auxiliary variables should be used to impute which analysis variables. In the imputation model, all analysis variables (exposure, mediators, outcomes, and confounders) were included in the imputation equation for every imputed variable. Additionally, we used tailored imputation equations to include only the auxiliary variables which were associated with the variable to be imputed in the regression models described above.

We used the Stata command -mi impute chained-with 10 cycles of regression switching and generated 40 imputed datasets. We imputed binary variables using binary logistic regression models, ordinal variables using ordinal logistic regression models, and all continuous variables using predicitve mean matching (with 10 nearest-neighbours) given evidence for a skewed distribution (Supplementary Table 2). Given that gang membership was a composite of two time points, gang membership at age 18 and at age 22 were imputed seperately and then combined to passively impute the composite measure of gang membership at either age, which was then included in the imputation equations for all analysis variables. After data were imputed, we compared the summary statistics (mean or proportion) for each analysis variable across complete case and imputed data to identify if there were any large differences. Convergence plots were also used to ensure that 10 cycles of regression switching was sufficient, and Monte Carlo errors were examined in the regression models for the outcomes and mediators to ensure that 40 imputed datasets were sufficient^32^.

After imputation, all analyses were combined across the 40 datasets using Rubin’s rules. For descriptive statistics and regression models, this was done using the Stata command -mi estimate-. Mediation models were estimated across each imputed dataset using a loop, with the results from each imputed dataset saved into a logfile. The log odds ratios and bootstrapped standard errors for the mediation effects (total causal effect, natural indirect effect, and natural direct effect) across each imputed dataset were extracted from the logfile. The mean of the log odds ratio for each mediation effect across the 40 imputed datasets was calculated, and the standard error was calculated using Rubin’s rules. Imputation and subsequent analysis code for the mediation models can be found on GitHub: https://github.com/gemmahammerton/gformula_1993_Pelotas

**Table S2.**
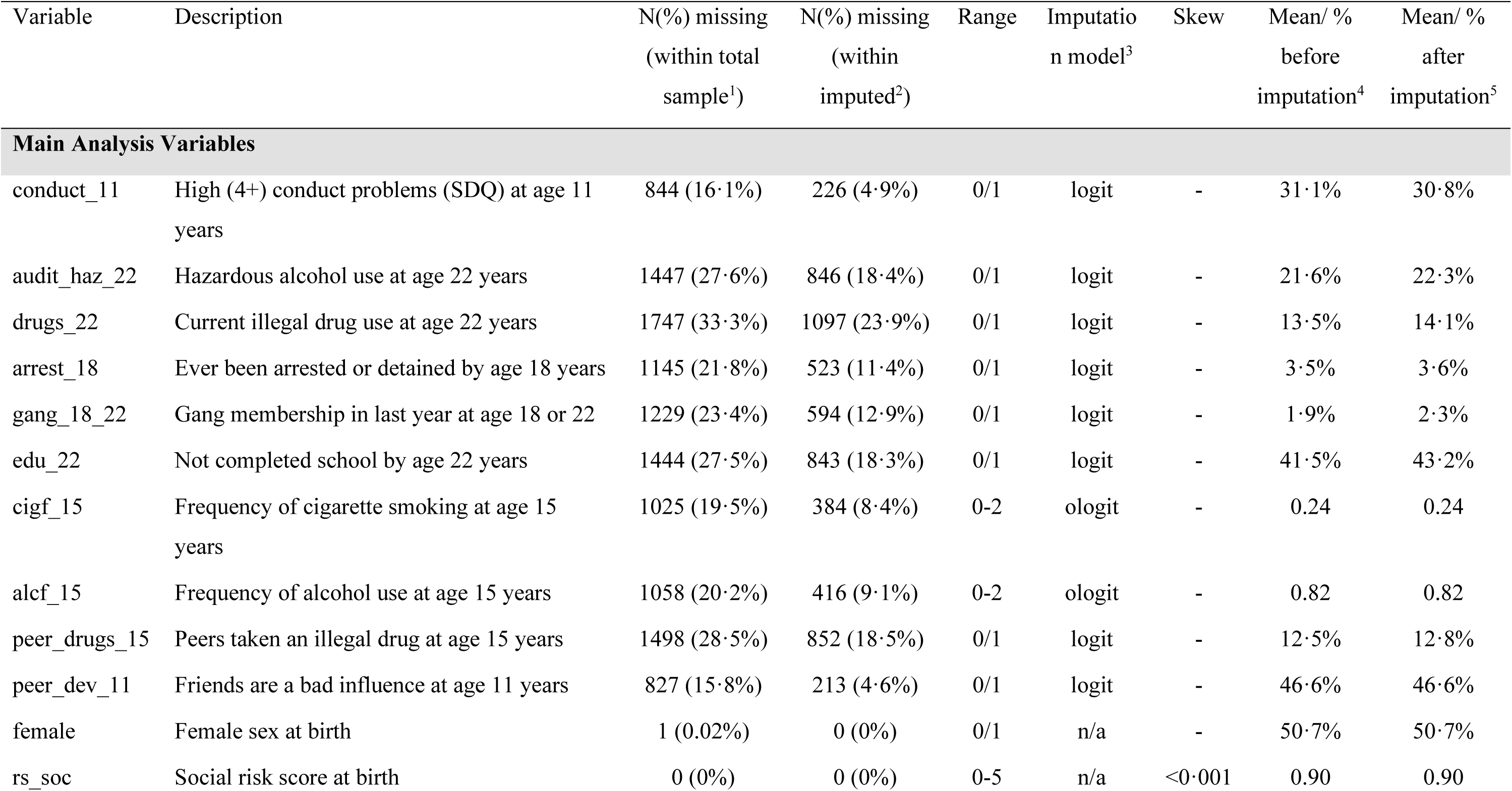
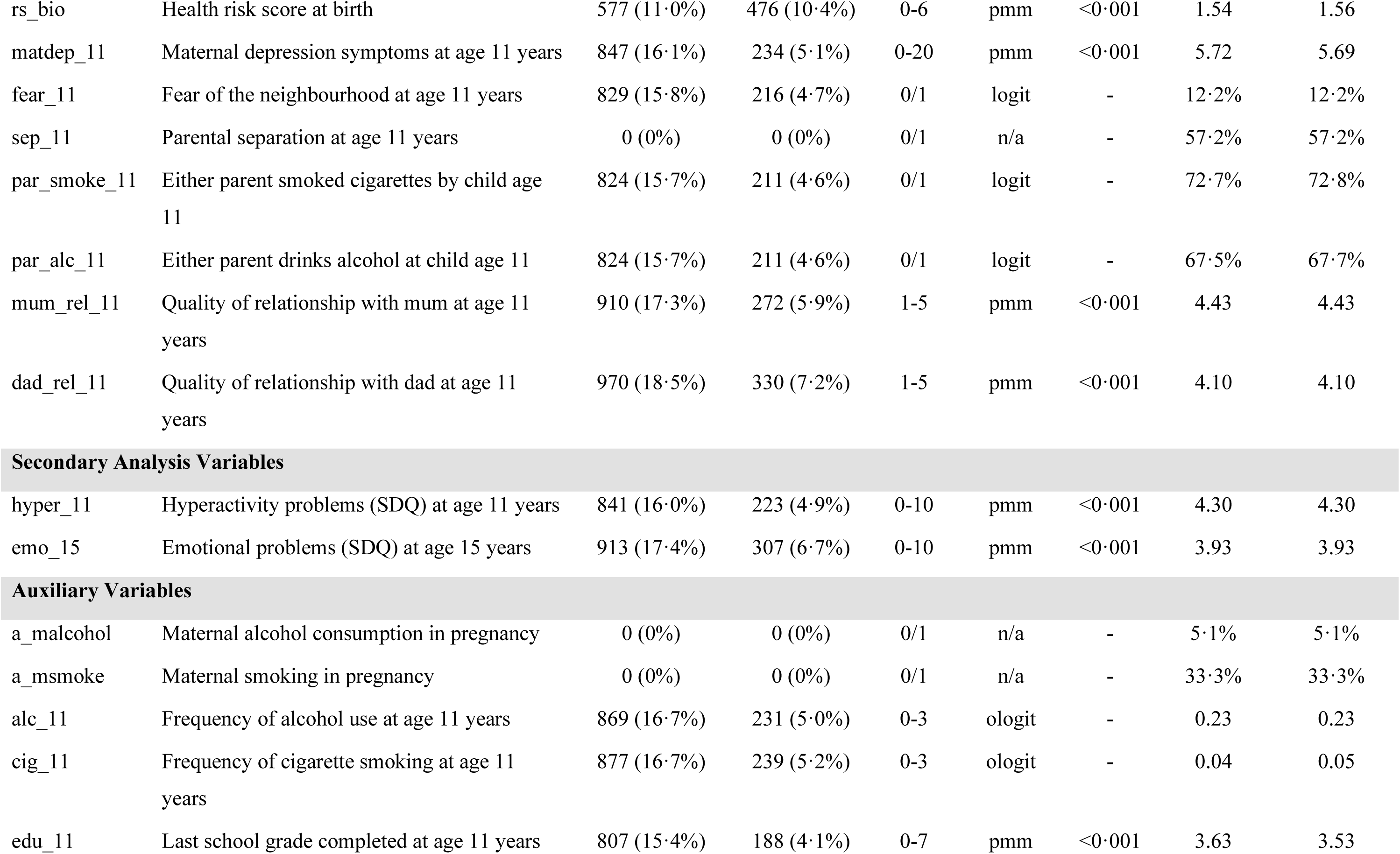
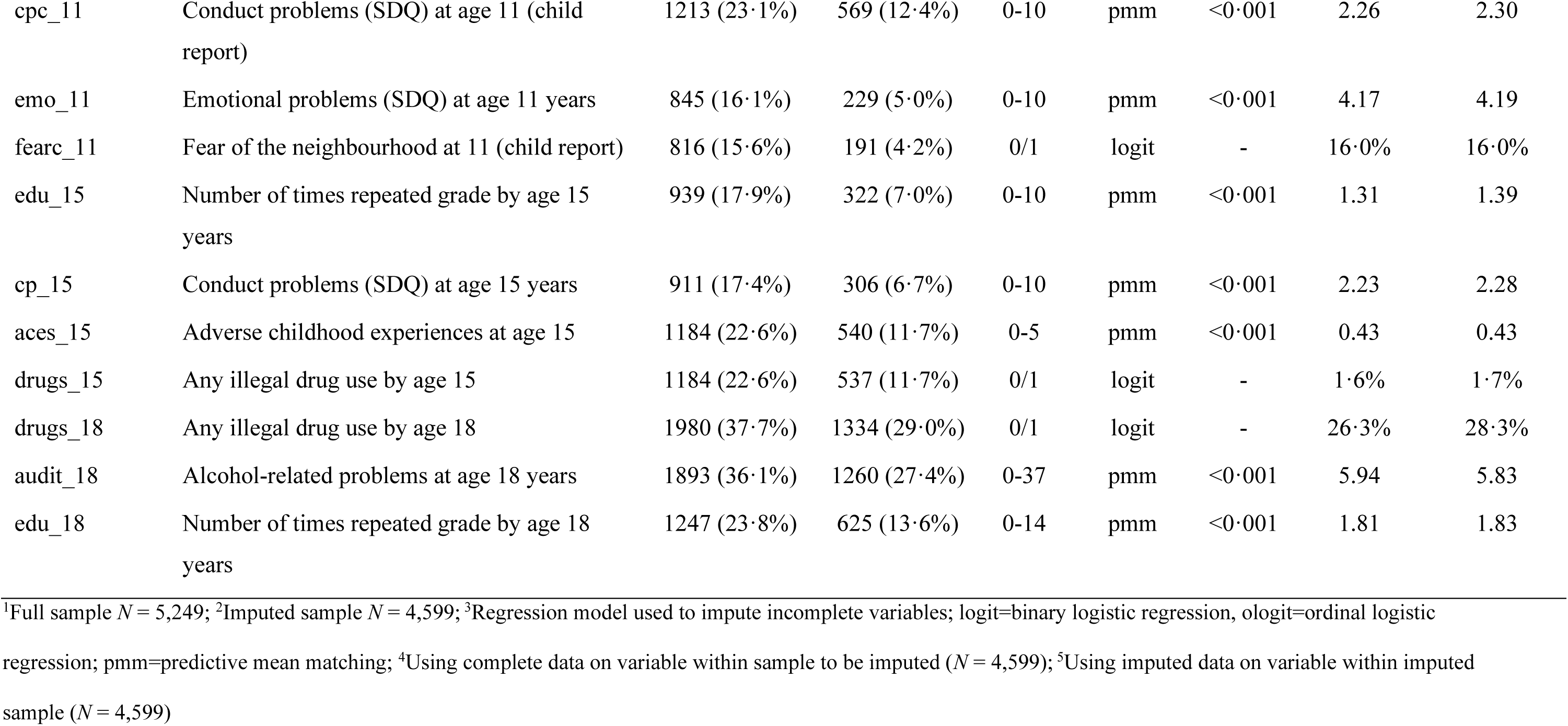
Summary of missing data for analysis and auxiliary variables.

| Variable | Description | N(%) missing<br>(within total<br>sample<sup>1</sup>) | N(%) missing<br>(within<br>imputed<sup>2</sup>) | Range | Imputatio<br>n model<sup>3</sup> | Skew | Mean/ %<br>before<br>imputation<sup>4</sup> | Mean/ %<br>after<br>imputation<sup>5</sup> |
| --- | --- | --- | --- | --- | --- | --- | --- | --- |
| **Main Analysis Variables** |  |  |  |  |  |  |  |  |
| conduct_11 | High (4+) conduct problems (SDQ) at age 11 years | 844 (16.1%) | 226 (4.9%) | 0/1 | logit | - | 31.1% | 30.8% |
| audit_haz_22 | Hazardous alcohol use at age 22 years | 1447 (27.6%) | 846 (18.4%) | 0/1 | logit | - | 21.6% | 22.3% |
| drugs_22 | Current illegal drug use at age 22 years | 1747 (33.3%) | 1097 (23.9%) | 0/1 | logit | - | 13.5% | 14.1% |
| arrest_18 | Ever been arrested or detained by age 18 years | 1145 (21.8%) | 523 (11.4%) | 0/1 | logit | - | 3.5% | 3.6% |
| gang_18_22 | Gang membership in last year at age 18 or 22 | 1229 (23.4%) | 594 (12.9%) | 0/1 | logit | - | 1.9% | 2.3% |
| edu_22 | Not completed school by age 22 years | 1444 (27.5%) | 843 (18.3%) | 0/1 | logit | - | 41.5% | 43.2% |
| cigf_15 | Frequency of cigarette smoking at age 15 years | 1025 (19.5%) | 384 (8.4%) | 0-2 | ologit | - | 0.24 | 0.24 |
| alcf_15 | Frequency of alcohol use at age 15 years | 1058 (20.2%) | 416 (9.1%) | 0-2 | ologit | - | 0.82 | 0.82 |
| peer_drugs_15 | Peers taken an illegal drug at age 15 years | 1498 (28.5%) | 852 (18.5%) | 0/1 | logit | - | 12.5% | 12.8% |
| peer_dev_11 | Friends are a bad influence at age 11 years | 827 (15.8%) | 213 (4.6%) | 0/1 | logit | - | 46.6% | 46.6% |
| female | Female sex at birth | 1 (0.02%) | 0 (0%) | 0/1 | n/a | - | 50.7% | 50.7% |
| rs_soc | Social risk score at birth | 0 (0%) | 0 (0%) | 0-5 | n/a | <0.001 | 0.90 | 0.90 |
| rs_bio | Health risk score at birth | 577 (11.0%) | 476 (10.4%) | 0-6 | pmm | <0.001 | 1.54 | 1.56 |
| matdep_11 | Maternal depression symptoms at age 11 years | 847 (16.1%) | 234 (5.1%) | 0-20 | pmm | <0.001 | 5.72 | 5.69 |
| fear_11 | Fear of the neighbourhood at age 11 years | 829 (15.8%) | 216 (4.7%) | 0/1 | logit | - | 12.2% | 12.2% |
| sep_11 | Parental separation at age 11 years | 0 (0%) | 0 (0%) | 0/1 | n/a | - | 57.2% | 57.2% |
| par_smoke_11 | Either parent smoked cigarettes by child age 11 | 824 (15.7%) | 211 (4.6%) | 0/1 | logit | - | 72.7% | 72.8% |
| par_alc_11 | Either parent drinks alcohol at child age 11 | 824 (15.7%) | 211 (4.6%) | 0/1 | logit | - | 67.5% | 67.7% |
| mum_rel_11 | Quality of relationship with mum at age 11 years | 910 (17.3%) | 272 (5.9%) | 1-5 | pmm | <0.001 | 4.43 | 4.43 |
| dad_rel_11 | Quality of relationship with dad at age 11 years | 970 (18.5%) | 330 (7.2%) | 1-5 | pmm | <0.001 | 4.10 | 4.10 |
| Secondary Analysis Variables |  |  |  |  |  |  |  |  |
| hyper_11 | Hyperactivity problems (SDQ) at age 11 years | 841 (16.0%) | 223 (4.9%) | 0-10 | pmm | <0.001 | 4.30 | 4.30 |
| emo_15 | Emotional problems (SDQ) at age 15 years | 913 (17.4%) | 307 (6.7%) | 0-10 | pmm | <0.001 | 3.93 | 3.93 |
| Auxiliary Variables |  |  |  |  |  |  |  |  |
| a_malcohol | Maternal alcohol consumption in pregnancy | 0 (0%) | 0 (0%) | 0/1 | n/a | - | 5.1% | 5.1% |
| a_msmoke | Maternal smoking in pregnancy | 0 (0%) | 0 (0%) | 0/1 | n/a | - | 33.3% | 33.3% |
| alc_11 | Frequency of alcohol use at age 11 years | 869 (16.7%) | 231 (5.0%) | 0-3 | ologit | - | 0.23 | 0.23 |
| cig_11 | Frequency of cigarette smoking at age 11 years | 877 (16.7%) | 239 (5.2%) | 0-3 | ologit | - | 0.04 | 0.05 |
| edu_11 | Last school grade completed at age 11 years | 807 (15.4%) | 188 (4.1%) | 0-7 | pmm | <0.001 | 3.63 | 3.53 |
| cpc_11 | Conduct problems (SDQ) at age 11 (child report) | 1213 (23.1%) | 569 (12.4%) | 0-10 | pmm | <0.001 | 2.26 | 2.30 |
| emo_11 | Emotional problems (SDQ) at age 11 years | 845 (16.1%) | 229 (5.0%) | 0-10 | pmm | <0.001 | 4.17 | 4.19 |
| fearc_11 | Fear of the neighbourhood at 11 (child report) | 816 (15.6%) | 191 (4.2%) | 0/1 | logit | - | 16.0% | 16.0% |
| edu_15 | Number of times repeated grade by age 15 years | 939 (17.9%) | 322 (7.0%) | 0-10 | pmm | <0.001 | 1.31 | 1.39 |
| cp_15 | Conduct problems (SDQ) at age 15 years | 911 (17.4%) | 306 (6.7%) | 0-10 | pmm | <0.001 | 2.23 | 2.28 |
| aces_15 | Adverse childhood experiences at age 15 | 1184 (22.6%) | 540 (11.7%) | 0-5 | pmm | <0.001 | 0.43 | 0.43 |
| drugs_15 | Any illegal drug use by age 15 | 1184 (22.6%) | 537 (11.7%) | 0/1 | logit | - | 1.6% | 1.7% |
| drugs_18 | Any illegal drug use by age 18 | 1980 (37.7%) | 1334 (29.0%) | 0/1 | logit | - | 26.3% | 28.3% |
| audit_18 | Alcohol-related problems at age 18 years | 1893 (36.1%) | 1260 (27.4%) | 0-37 | pmm | <0.001 | 5.94 | 5.83 |
| edu_18 | Number of times repeated grade by age 18 years | 1247 (23.8%) | 625 (13.6%) | 0-14 | pmm | <0.001 | 1.81 | 1.83 |

<sup>1</sup>Full sample $N = 5,249$ ; <sup>2</sup>Imputed sample $N = 4,599$ ; <sup>3</sup>Regression model used to impute incomplete variables; logit=binary logistic regression, ologit=ordinal logistic
regression; pmm=predictive mean matching; <sup>4</sup>Using complete data on variable within sample to be imputed ( $N = 4,599$ ); <sup>5</sup>Using imputed data on variable within imputed sample ( $N = 4,599$ )

**Table S3.** Univariable associations for baseline confounders with mediators and outcomes using imputed data; *N* = 4,599.

| OR (95% CI); $p$ value | Mediators | | | Outcomes | |
| --- | --- | --- | --- | --- | --- |
|  | Police arrest | Gang membership | School non-completion | Hazardous alcohol use | Illicit drug use |
| **Baseline confounders** |  |  |  |  |  |
| Female sex | 0.14 (0.09, 0.23);<br><0.001 | 0.19 (0.10, 0.35);<br><0.001 | 0.57 (0.50, 0.64);<br><0.001 | 0.38 (0.33, 0.45);<br><0.001 | 0.42 (0.34, 0.51);<br><0.001 |
| Sociodemographic risk score | 1.25 (1.06, 1.47);<br>0.007 | 1.42 (1.16, 1.75);<br>0.001 | 2.23 (2.07, 2.40);<br><0.001 | 0.93 (0.86, 1.01);<br>0.082 | 0.91 (0.83, 1.01);<br>0.069 |
| Health risk score | 1.19 (1.02, 1.40);<br>0.030 | 1.15 (0.93, 1.43);<br>0.189 | 1.50 (1.41, 1.60);<br><0.001 | 1.02 (0.95, 1.09);<br>0.620 | 1.00 (0.91, 1.09);<br>0.960 |
| Maternal depression | 1.06 (1.03, 1.10);<br>0.001 | 1.09 (1.05, 1.14);<br><0.001 | 1.10 (1.08, 1.12);<br><0.001 | 1.00 (0.98, 1.02);<br>0.807 | 1.00 (0.98, 1.03);<br>0.720 |
| Fear of the neighbourhood | 1.21 (0.75, 1.95);<br>0.424 | 1.37 (0.74, 2.56);<br>0.317 | 1.51 (1.25, 1.83);<br><0.001 | 0.81 (0.63, 1.04);<br>0.101 | 1.25 (0.96, 1.65);<br>0.101 |
| Parental separation | 2.39 (1.62, 3.53);<br><0.001 | 2.24 (1.37, 3.66);<br>0.001 | 2.95 (2.59, 3.36);<br><0.001 | 1.12 (0.95, 1.31);<br>0.174 | 1.17 (0.97, 1.42);<br>0.107 |
| Parental smoking | 1.61 (1.07, 2.44);<br>0.024 | 1.12 (0.68, 1.85);<br>0.666 | 1.70 (1.46, 1.97);<br><0.001 | 1.07 (0.89, 1.28);<br>0.455 | 1.03 (0.83, 1.28);<br>0.760 |
| Parental alcohol use | 1.02 (0.71, 1.46);<br>0.908 | 0.63 (0.40, 1.00);<br>0.051 | 0.87 (0.76, 1.00);<br>0.050 | 1.37 (1.16, 1.63);<br><0.001 | 1.32 (1.07, 1.64);<br>0.010 |
| Father-child relationship | 0.84 (0.74, 0.95);<br>0.005 | 0.82 (0.69, 0.97);<br>0.022 | 0.74 (0.70, 0.79);<br><0.001 | 0.96 (0.89, 1.02);<br>0.191 | 0.91 (0.84, 0.99);<br>0.024 |
| Mother-child relationship | 0.94 (0.78, 1.12);<br>0.469 | 0.65 (0.53, 0.78);<br><0.001 | 0.61 (0.57, 0.66);<br><0.001 | 0.91 (0.83, 0.99);<br>0.028 | 0.98 (0.87, 1.11);<br>0.764 |
| Hyperactivity problems | 1.15 (1.09, 1.21);<br><0.001 | 1.26 (1.17, 1.36);<br><0.001 | 1.20 (1.18, 1.23);<br><0.001 | 1.08 (1.06, 1.11);<br><0.001 | 1.04 (1.01, 1.07);<br>0.010 |

OR = Odds Ratio, 95% CI = 95% Confidence Interval

**Table S4.** Univariable associations for intermediate confounders with mediators and outcomes using imputed data; *N* = 4,599.

| OR (95% CI);<br>*p* value | Police arrest | Gang membership | School non-completion | Hazardous<br>alcohol use | Illicit drug use |
| --- | --- | --- | --- | --- | --- |
| **Intermediate confounders** |  |  |  |  |  |
| Peer deviance | 2.23 (1.56, 3.19);<br><0.001 | 2.91 (1.75, 4.86);<br><0.001 | 1.57 (1.38, 1.79);<br><0.001 | 1.39 (1.20, 1.62);<br><0.001 | 1.67 (1.36, 2.04);<br><0.001 |
| Frequency of cigarette smoking (never) | Reference |  |  |  |  |
| Not in last month | 2.22 (1.40, 3.53) | 1.46 (0.76, 2.81) | 1.69 (1.39, 2.04) | 1.07 (0.84, 1.36) | 1.81 (1.40, 2.36) |
| 1 to 5 days in last month | 3.81 (2.33, 6.22) | 2.45 (1.17, 5.11) | 3.29 (2.44, 4.45) | 1.78 (1.32, 2.40) | 1.89 (1.30, 2.75) |
| Frequency of alcohol use (never) | Reference |  |  |  |  |
| Not in last month | 1.78 (1.13, 2.80) | 1.27 (0.72, 2.26) | 0.69 (0.59, 0.80) | 1.45 (1.19, 1.75) | 2.20 (1.69, 2.86) |
| 1 to 5 days in last month | 2.30 (1.44, 3.67) | 1.61 (0.90, 2.87) | 0.84 (0.72, 0.99) | 2.00 (1.63, 2.45) | 3.12 (2.37, 4.09) |
| Peer drug use | 2.43 (1.57, 3.76);<br><0.001 | 2.47 (1.40, 4.38);<br>0.002 | 1.42 (1.14, 1.77);<br>0.002 | 1.39 (1.10, 1.76);<br>0.006 | 2.53 (1.97, 3.27);<br><0.001 |
| Emotional problems | 0.99 (0.93, 1.05);<br>0.700 | 1.02 (0.94, 1.10);<br>0.704 | 1.10 (1.07, 1.13);<br><0.001 | 0.97 (0.94, 1.00);<br>0.060 | 0.95 (0.92, 0.99);<br>0.010 |

OR = Odds Ratio, 95% CI = 95% Confidence Interval

**Table S5.**
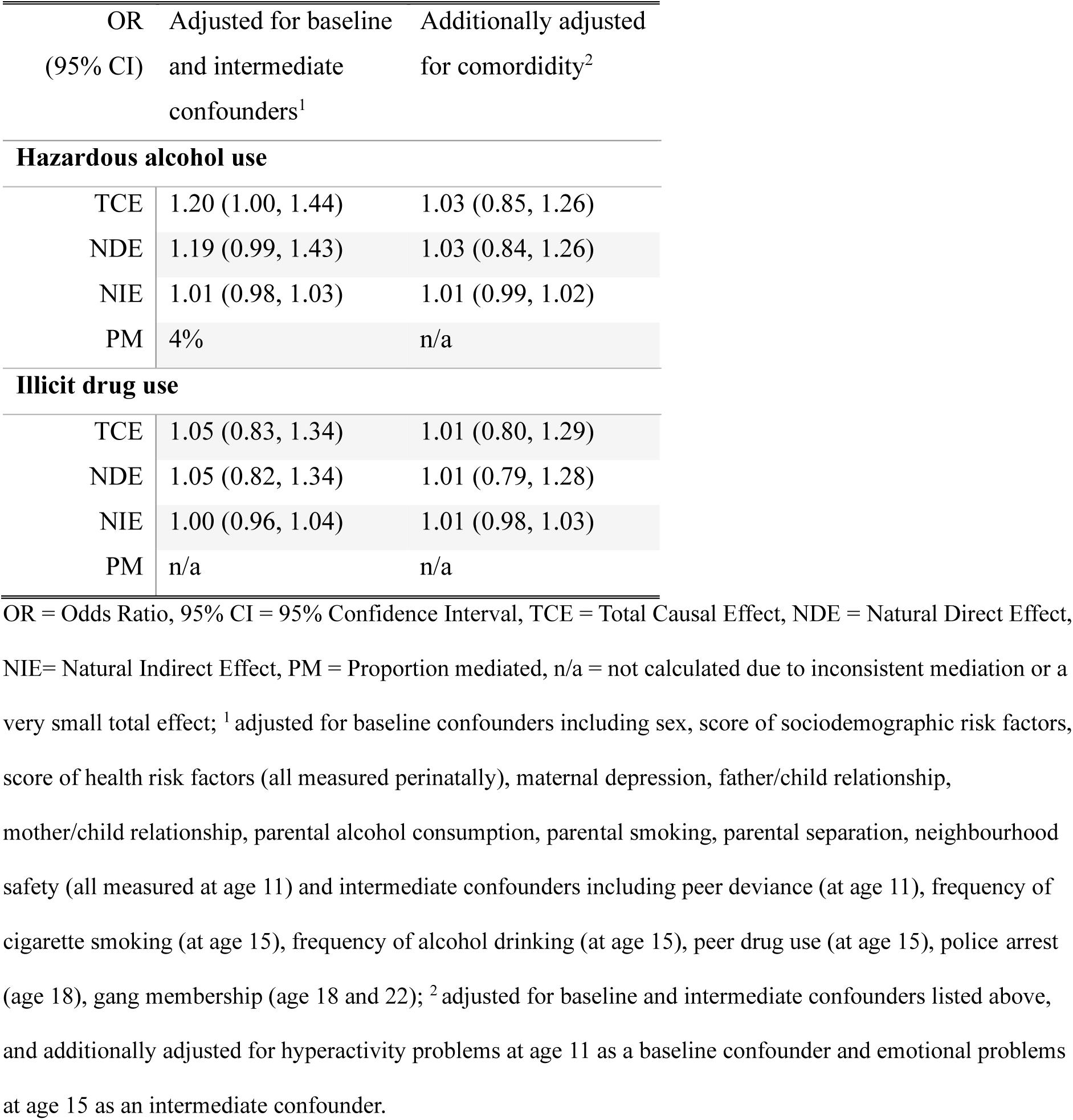
Mediation models for school non-completion with police arrest and gang membership treated as intermediate confounders using imputed data; *N* = 4,599.

| OR<br>(95% CI) | Adjusted for baseline<br>and intermediate<br>confounders<sup>1</sup> | Additionally adjusted<br>for comorbidity<sup>2</sup> |
| --- | --- | --- |
| **Hazardous alcohol use** |  |  |
| TCE | 1.20 (1.00, 1.44) | 1.03 (0.85, 1.26) |
| NDE | 1.19 (0.99, 1.43) | 1.03 (0.84, 1.26) |
| NIE | 1.01 (0.98, 1.03) | 1.01 (0.99, 1.02) |
| PM | 4% | n/a |
| **Illicit drug use** |  |  |
| TCE | 1.05 (0.83, 1.34) | 1.01 (0.80, 1.29) |
| NDE | 1.05 (0.82, 1.34) | 1.01 (0.79, 1.28) |
| NIE | 1.00 (0.96, 1.04) | 1.01 (0.98, 1.03) |
| PM | n/a | n/a |

OR = Odds Ratio, 95% CI = 95% Confidence Interval, TCE = Total Causal Effect, NDE = Natural Direct Effect, NIE = Natural Indirect Effect, PM = Proportion mediated, n/a = not calculated due to inconsistent mediation or a very small total effect; <sup>1</sup> adjusted for baseline confounders including sex, score of sociodemographic risk factors, score of health risk factors (all measured perinatally), maternal depression, father/child relationship, mother/child relationship, parental alcohol consumption, parental smoking, parental separation, neighbourhood safety (all measured at age 11) and intermediate confounders including peer deviance (at age 11), frequency of cigarette smoking (at age 15), frequency of alcohol drinking (at age 15), peer drug use (at age 15), police arrest (age 18), gang membership (age 18 and 22); <sup>2</sup> adjusted for baseline and intermediate confounders listed above, and additionally adjusted for hyperactivity problems at age 11 as a baseline confounder and emotional problems at age 15 as an intermediate confounder.

**Table S6.** Associations between the exposure (conduct problems at age 11 years) and the mediators (police arrest, gang membership, and school non-completion between age 18 and 22 years) using complete case data; N = 2,342.

| OR (95% CI);<br>*p* value | Unadjusted | Adjusted for baseline<br>confounders<sup>1</sup> | Additionally adjusted<br>for hyperactivity<sup>2</sup> |
| --- | --- | --- | --- |
| Police arrest | 1.91 (1.14, 3.17);<br>0.013 | 1.47 (0.85, 2.53);<br>0.166 | 1.16 (0.65, 2.08);<br>0.617 |
| Gang membership | 2.55 (1.31, 4.98);<br>0.006 | 1.78 (0.87, 3.64);<br>0.112 | 1.25 (0.58, 2.70);<br>0.561 |
| School non-completion | 2.64 (2.19, 3.18);<br><0.001 | 1.89 (1.53, 2.34);<br><0.001 | 1.54 (1.23, 1.94);<br><0.001 |

OR = Odds Ratio, 95% CI = 95% Confidence Interval; <sup>1</sup> adjusted for baseline confounders including sex, score of sociodemographic risk factors, score of health risk factors (all measured perinatally), maternal depression, father/child relationship, mother/child relationship, parental alcohol consumption, parental smoking, parental separation, neighbourhood safety (all measured at age 11); <sup>2</sup> adjusted for baseline confounders listed above, and additionally adjusted for hyperactivity problems at age 11

**Table S7.**
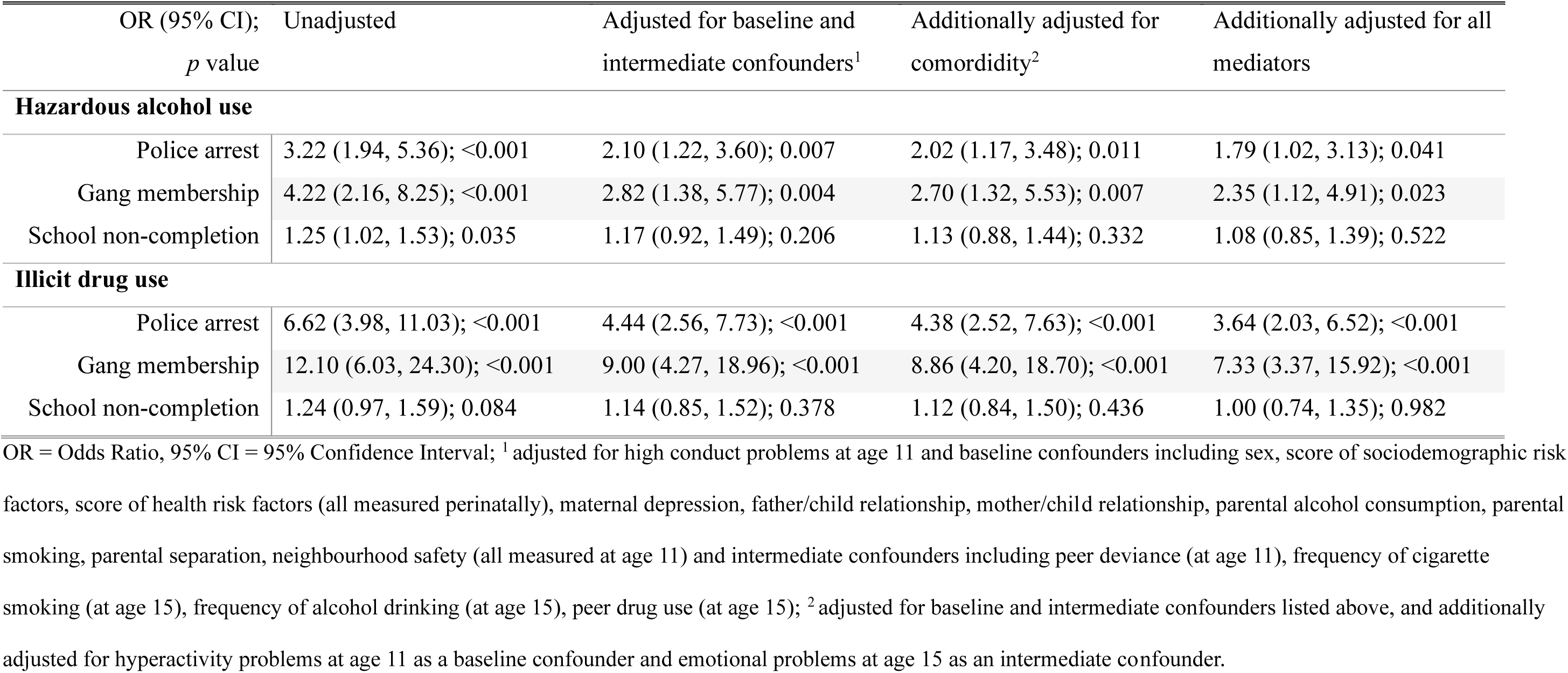
Associations between the mediators (police arrest, gang membership, and school non-completion between age 18 and 22 years) and the outcomes (hazardous alcohol use and illicit drug use at age 22 years) using complete case data; N = 2,342.

| OR (95% CI);<br>*p* value | Unadjusted | Adjusted for baseline and<br>intermediate confounders<sup>1</sup> | Additionally adjusted for<br>comorbidity<sup>2</sup> | Additionally adjusted for all<br>mediators |
| --- | --- | --- | --- | --- |
| **Hazardous alcohol use** |  |  |  |  |
| Police arrest | 3.22 (1.94, 5.36); <0.001 | 2.10 (1.22, 3.60); 0.007 | 2.02 (1.17, 3.48); 0.011 | 1.79 (1.02, 3.13); 0.041 |
| Gang membership | 4.22 (2.16, 8.25); <0.001 | 2.82 (1.38, 5.77); 0.004 | 2.70 (1.32, 5.53); 0.007 | 2.35 (1.12, 4.91); 0.023 |
| School non-completion | 1.25 (1.02, 1.53); 0.035 | 1.17 (0.92, 1.49); 0.206 | 1.13 (0.88, 1.44); 0.332 | 1.08 (0.85, 1.39); 0.522 |
| **Illicit drug use** |  |  |  |  |
| Police arrest | 6.62 (3.98, 11.03); <0.001 | 4.44 (2.56, 7.73); <0.001 | 4.38 (2.52, 7.63); <0.001 | 3.64 (2.03, 6.52); <0.001 |
| Gang membership | 12.10 (6.03, 24.30); <0.001 | 9.00 (4.27, 18.96); <0.001 | 8.86 (4.20, 18.70); <0.001 | 7.33 (3.37, 15.92); <0.001 |
| School non-completion | 1.24 (0.97, 1.59); 0.084 | 1.14 (0.85, 1.52); 0.378 | 1.12 (0.84, 1.50); 0.436 | 1.00 (0.74, 1.35); 0.982 |

OR = Odds Ratio, 95% CI = 95% Confidence Interval; <sup>1</sup> adjusted for high conduct problems at age 11 and baseline confounders including sex, score of sociodemographic risk factors, score of health risk factors (all measured perinatally), maternal depression, father/child relationship, mother/child relationship, parental alcohol consumption, parental smoking, parental separation, neighbourhood safety (all measured at age 11) and intermediate confounders including peer deviance (at age 11), frequency of cigarette smoking (at age 15), frequency of alcohol drinking (at age 15), peer drug use (at age 15); <sup>2</sup> adjusted for baseline and intermediate confounders listed above, and additionally adjusted for hyperactivity problems at age 11 as a baseline confounder and emotional problems at age 15 as an intermediate confounder.

**Table S8.** Mediation models including all mediators (police arrest, gang membership, and school non-completion) simultaneously using complete case data; N = 2,342.

| OR<br>(95% CI) | Unadjusted | Adjusted for<br>baseline and<br>intermediate<br>confounders<sup>1</sup> | Additionally<br>adjusted for<br>comorbidity<sup>2</sup> |
| --- | --- | --- | --- |
| **Hazardous alcohol use** |  |  |  |
| TCE | 1.29 (1.04, 1.61) | 1.23 (1.00, 1.53) | 1.08 (0.82, 1.42) |
| NDE | 1.22 (0.97, 1.52) | 1.19 (0.95, 1.49) | 1.06 (0.80, 1.40) |
| NIE | 1.06 (1.00, 1.13) | 1.04 (0.99, 1.08) | 1.02 (0.99, 1.05) |
| PM | 24% | 17% | 22% |
| **Illicit drug use** |  |  |  |
| TCE | 1.18 (0.95, 1.48) | 1.07 (0.82, 1.40) | 0.99 (0.73, 1.35) |
| NDE | 1.08 (0.85, 1.36) | 1.01 (0.76, 1.34) | 0.97 (0.71, 1.32) |
| NIE | 1.10 (0.99, 1.22) | 1.06 (0.99, 1.14) | 1.02 (0.96, 1.10) |
| PM | 57% | 87% | n/a |

OR = Odds Ratio, 95% CI = 95% Confidence Interval, TCE = Total Causal Effect, NDE = Natural Direct Effect, NIE = Natural Indirect Effect, PM = Proportion mediated, n/a = not calculated due to inconsistent mediation; <sup>1</sup> adjusted for baseline confounders including sex, score of sociodemographic risk factors, score of health risk factors (all measured perinatally), maternal depression, mother/child relationship, father/child relationship, parental alcohol consumption, parental smoking, parental separation, neighbourhood safety (all measured at age 11) and intermediate confounders including peer deviance (at age 11), frequency of cigarette smoking (at age 15), frequency of alcohol drinking (at age 15), peer drug use (at age 15); <sup>2</sup>adjusted for baseline and intermediate confounders listed above, and additionally adjusted for hyperactivity problems at age 11 as a baseline confounder and emotional problems at age 15 as an intermediate confounder.

